# Symptoms and SARS-CoV-2 positivity in the general population in the UK

**DOI:** 10.1101/2021.08.19.21262231

**Authors:** Karina-Doris Vihta, Koen B. Pouwels, Tim Peto, Emma Pritchard, David W. Eyre, Thomas House, Owen Gethings, Ruth Studley, Emma Rourke, Duncan Cook, Ian Diamond, Derrick Crook, Philippa C. Matthews, Nicole Stoesser, Ann Sarah Walker, the COVID-19 Infection Survey team

## Abstract

**Background:** Several community-based studies have assessed the ability of different symptoms to identify COVID-19 infections, but few have compared symptoms over time (reflecting SARS-CoV-2 variants) and by vaccination status.

**Methods:** Using data and samples collected by the COVID-19 Infection Survey at regular visits to representative households across the UK, we compared symptoms in new PCR-positives and comparator test-negative controls.

**Results:** From 26/4/2020-7/8/2021, 27,869 SARS-CoV-2 PCR-positive episodes occurred in 27,692 participants (median 42 years (IQR 22-58)); 13,427 (48%) self-reported symptoms (“symptomatic positive episodes”). The comparator group comprised 3,806,692 test-negative visits (457,215 participants); 130,612 (3%) self-reported symptoms (“symptomatic negative visit”). Reporting of any symptoms in positive episodes varied over calendar time, reflecting changes in prevalence of variants, incidental changes (e.g. seasonal pathogens, schools re-opening) and vaccination roll-out. There was a small increase in sore throat reporting in symptomatic positive episodes and negative visits from April-2021. After May-2021 when Delta emerged there were substantial increases in headache and fever in positives, but not in negatives. Although specific symptom reporting in symptomatic positive episodes vs. negative visits varied by age, sex, and ethnicity, only small improvements in symptom-based infection detection were obtained; e.g. adding fatigue/weakness or all eight symptoms to the classic four symptoms (cough, fever, loss of taste/smell) increased sensitivity from 74% to 81% to 90% but tests per positive from 4.6 to 5.3 to 8.7.

**Conclusions:** Whilst SARS-CoV-2-associated symptoms vary by variant, vaccination status and demographics, differences are modest and do not warrant large-scale changes to targeted testing approaches given resource implications.

**Summary:** Within the COVID-19 Infection Survey, recruiting representative households across the UK general population, SARS-CoV-2-associated symptoms varied by viral variant, vaccination status and demographics. However, differences are modest and do not currently warrant large-scale changes to targeted testing approaches.

## INTRODUCTION

Whilst a substantial proportion of individuals infected with SARS-CoV-2 remain asymptomatic[1,2], symptoms are associated with higher viral loads[3], and higher viral loads with infectivity and transmission[4]. Resource constraints generally prevent universal testing being deployed, so testing and isolation strategies are usually targeted to those with symptoms most predictive of infection and/or contacts of known positive individuals. Currently, four “classic” symptoms trigger PCR-based community testing in the UK, namely: loss/change of smell/taste, fever, and/or a new, continuous cough. In the US, the Centers for Disease Control and Prevention (CDC) advises testing for any of fever or chills, cough, shortness of breath/difficulty breathing, new loss of taste/smell, fatigue, muscle or body aches, headache, sore throat, congestion/runny nose, nausea/vomiting, or diarrhoea.

As national testing policies depend on symptoms, understanding their predictive value in the context of seasonality, changing prevalence of different variants[3], and widespread vaccination[5] is essential. Most studies to date have restricted to those hospitalised or seeking healthcare, who do not represent most infections[6]. Three recent UK community-based studies suggested that sensitivity could be increased by 10-20% by extending the “classic” symptoms (REACT[7], adding combinations of headache, muscle aches, chills and appetite loss depending on age; ZOE[8], adding different symptoms depending on age, sex, BMI and working in healthcare; VirusWatch[9], adding feeling feverish, headache, muscle aches, loss of appetite or chills) but at a cost of increasing numbers eligible for testing 2-3 fold and tests per positive identified up to 7-fold. However, these studies were mainly before widespread vaccination, and whilst Alpha dominated. ZOE found no evidence of difference in reported symptoms between wild type and the Alpha variant[10]. With Delta, ZOE has recently identified headache, sore throat and runny nose/sneezing as non-classic symptoms most commonly occurring in fully, partially and unvaccinated positives, respectively[11].

We therefore used a large community-based survey representative of the UK general population to investigate symptoms over time in PCR-positive episodes and negative controls, also evaluating the impact of age, ethnicity, cycle threshold (Ct) value, vaccination status and PCR gene profile (as a proxy for variant).

## METHODS

The Office for National Statistics (ONS) COVID-19 Infection Survey[12] (ISRCTN21086382, https://www.ndm.ox.ac.uk/covid-19/covid-19-infection-survey/protocol-and-information-sheets) continuously randomly selects private households from address lists and previous surveys. Having obtained verbal agreement, each household is visited by a study worker, and written informed consent obtained for individuals aged ≥2y (from parents/carers for those 2-15y, with those 10-15y also providing written assent). At the first visit, participants are asked for consent for optional follow-up visits every week for the next month, then monthly thereafter. At each visit, participants provide a nose and throat self-swab following instructions from the study worker and answer questions about behaviours, work, vaccination uptake and symptoms in the last 7 days (https://www.ndm.ox.ac.uk/covid-19/covid-19-infection-survey/case-record-forms). Twelve specific symptoms are elicited, namely: loss of taste, loss of smell, fever, cough, headache, tiredness/weakness (denoted fatigue/weakness), muscle ache (denoted muscle ache/myalgia), abdominal pain, diarrhoea, nausea or vomiting, shortness of breath and sore throat; plus a general question about presence of symptoms participants considered COVID-19-related (denoted any evidence of symptoms).

Swabs are tested at national testing laboratories using the Thermo Fisher TaqPath PCR assay (3 targets: ORF1ab, nucleocapsid (N), and spike protein (S)). If N and/or ORF1ab genes are detected, samples are called positive; the S-gene can accompany other genes, but does not count as positive alone (27/34,494 (0.08%) only S-gene positives counted as negative).

We included the first study positive test in each positive “episode”, defining re-infections (arbitrarily) as occurring ≥120 days after an index positive with a preceding negative test, or after 4 consecutive negative tests[5]. Each episode was classified by the minimum Ct value across positive tests in it and as wild-type/Delta-compatible if the S-gene was ever detected within it (by definition, with N/ORF1ab/both), otherwise Alpha-compatible if positive at least once for ORF1ab+N, otherwise “other” (N-only/ORF1ab-only) (**Fig.S1**). Symptom presence included reports at any visit (test-positive/negative/failed) within [0,+35] days of the first positive per episode (i.e. spanning [-7,+35] days given the question timeframe).

The comparator was visits with negative PCR tests, excluding visits with symptoms related to ongoing COVID-19 (and long COVID), with high probability of undetected COVID-19, and where symptoms were likely driven by recent vaccination (details in Supplementary Methods).

### Statistical analyses

Primary analyses restricted to positive episodes and negative visits with evidence of any symptoms, because this is the population targeted by public health messages to test and self-isolate, denoted “symptomatic positive episodes” and “symptomatic negative visits” respectively. We considered all positive episodes, as well as subgroups defined by Ct (reflecting viral load), gene positivity pattern (reflecting variant compatibility), vaccination status, age and calendar time (reflecting background incidental symptoms) (details in Supplementary Methods).

Initially, hierarchical clustering with complete linkage (Jaccard distance matrix) assessed congruence of self-reported symptoms internally. To assess predictors of reporting any symptom, and each symptom in symptomatic episodes/visits, we fitted generalised additive models (binomial distribution with complementary log-log link, mgcv (v.1.8-31) package), adjusting simultaneously for calendar time (smoothing spline), age (smoothing spline), sex, ethnicity (white vs non-white). We tested whether effects varied by positive/negative episodes/visits using interaction tests. In positives only, separate models also adjusted for Ct value (smoothing spline) and gene-positivity, or vaccination status (details in Supplementary Methods).

We assessed the impact on performance of adding each of the other eight symptoms to the four classic symptoms currently prompting testing in the UK, and of every combination of 1-8 symptoms, and any of the 12 elicited symptoms, using standard metrics (details in Supplementary Methods, epiR (v.1.0-15), pROC (v.1.16.2) packages) plus tests per case (TPC)=1/positive predictive value (PPV) and the inflation factor=episodes/visits with any included symptom/episodes/visits with classic symptoms. Finally, for each positive episode, we compared symptoms reported at the first vs subsequent visits within the episode (both absent, both present, absent then present, present then absent), and the associated Ct distributions.

## RESULTS

Between 26/4/2020-7/8/2021, 5,130,318 study PCR results were available from 484,317 participants; 34,494 (0.67%) were SARS-CoV-2-positive. In total, 27,869 positive episodes occurred in 27,692 participants (median age 42 years (IQR 22-58)); 13,427 (48%) self-reported symptoms (“symptomatic positive episodes”). The comparator group comprised 3,806,692 negative visits (457,215 participants, median age 52 years (IQR 32-66)); 130,612 (3%) self-reported symptoms (“symptomatic negative visits”) (exclusions in **Tables S1&S2**; characteristics in **Table S3**).

### Specific symptoms are associated with SARS-CoV-2 and variants

Fatigue/weakness, cough, and headache were the most frequently reported symptoms in positive episodes (54%, 54%, 52% of symptomatic positive episodes; **Fig.1**). However, headache and cough were most frequently reported amongst negative visits too, together with sore throat (23%, 22%, 22%; **Fig.1**). Loss of taste/smell were the most specific symptoms for SARS-CoV-2 positivity (33%/33% positive episodes, only 2%/2% negative visits). In positive episodes, loss of taste or smell were commonly co-reported, as were gastrointestinal symptoms, and headache/myalgia/fatigue. Symptom co-reporting in symptomatic positive episodes was generally similar regardless of Ct, variant, vaccination status or age (**Fig.S2**), although myalgia was less strongly co-reported with headache/fatigue in those 6-15y, with Delta and in those ≥14d post-second vaccination. Patterns were broadly similar in negatives, except cough and sore throat were more commonly co-reported.

**Figure 1.**
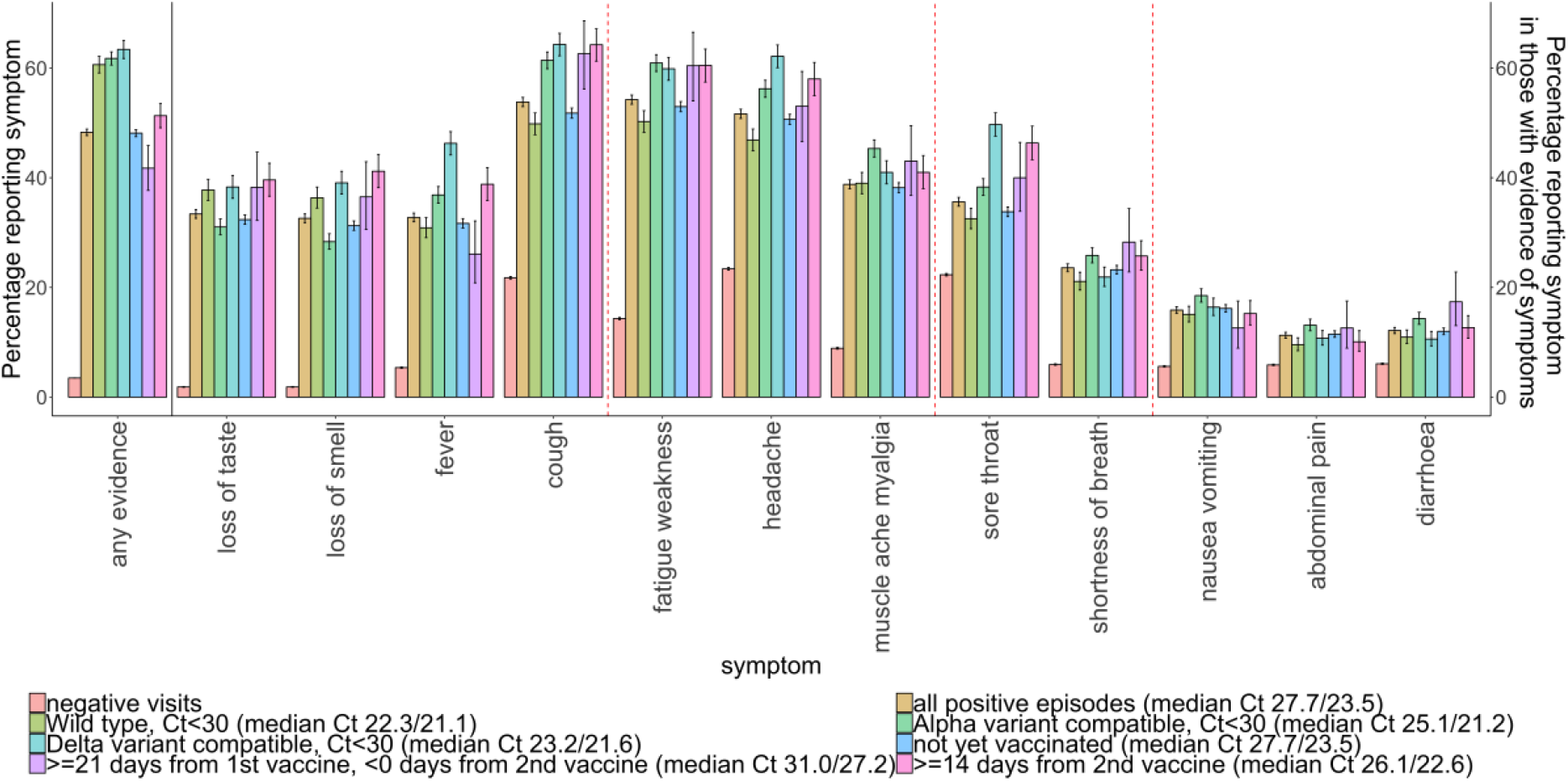
Percentage self-reporting any evidence of symptoms out of all positive episodes and negative visits, and percentage reporting each specific symptom of those reporting any symptoms Note: wild-type defined as S-gene positive before 17 November 2020; Alpha-compatible defined as S-gene negative from 17 November 2020 through 17 May 2021, Delta-compatible defined as S-gene positive from 17 May 2021. Post-vaccination positives split into not yet vaccinated, those 21 days after first vaccination and before second vaccination, and 14 days or more after second vaccination. The two median values are median Ct in all and symptomatic positive episodes in each group. See Fig.6 for associations between Ct and symptoms. Red dashed lines indicate symptom clusters based on hierarchical clustering (**Fig.S2**).

Symptomatology varied significantly by variant (**Fig.1**); with a smaller percentage of symptomatic positive episodes reporting loss of taste/smell for Alpha-compatible (31%/28%) than wild-type (38%/36%) or Delta-compatible (38%/39%) symptomatic positive episodes (p<0.0001). Fever/headache/sore throat had the largest difference between symptoms reported for Alpha-compatible (37%/56%/38%) and Delta-compatible (46%/62%/50%) symptomatic positive episodes (p<0.0001). Cough and fatigue/weakness had the largest differences between wild-type (50%/50%) vs Alpha-compatible (61%/61%) or Delta-compatible (64%/60%) symptomatic positive episodes (p<0.0001). In general, specific symptoms were reported slightly more in symptomatic positive episodes ≥14d from second vaccination versus those unvaccinated or ≥21d from first vaccination.

### Symptomatology over calendar time

Adjusting for age, sex and ethnicity, the probability of symptoms being reported amongst positive episodes was reasonably stable after August-2020 given changes in incidence and sample size (**Fig.2**, top panels), with fluctuations likely reflecting school return in September-2020 and March-2021, plus the emergence of Alpha and Delta in November-2020 and May-2021. Smaller fluctuations in reporting any symptoms in negative visits (**Fig.2**, bottom panels) mirrored those in positive episodes. The percentage of symptomatic positive episodes reporting each specific symptom generally increased over time, consistent with increasing awareness. Reporting of most specific symptoms except loss of taste/smell temporarily peaked in January-2021, consistent with the peak in Alpha, then remained approximately constant through to May-2021, before increasing again, markedly so for headache, cough and fever after Delta became dominant. Increases in cough and sore throat in symptomatic negative visits occurred after school return in September-2020, in January-2021 when schools were shut, and from early April-2021, consistent with other respiratory viruses. The winter months saw particular increases in gastrointestinal symptoms, fatigue/weakness, myalgia and headache in symptomatic negative visits, consistent with the presence of other common seasonal pathogens.

**Figure 2.**
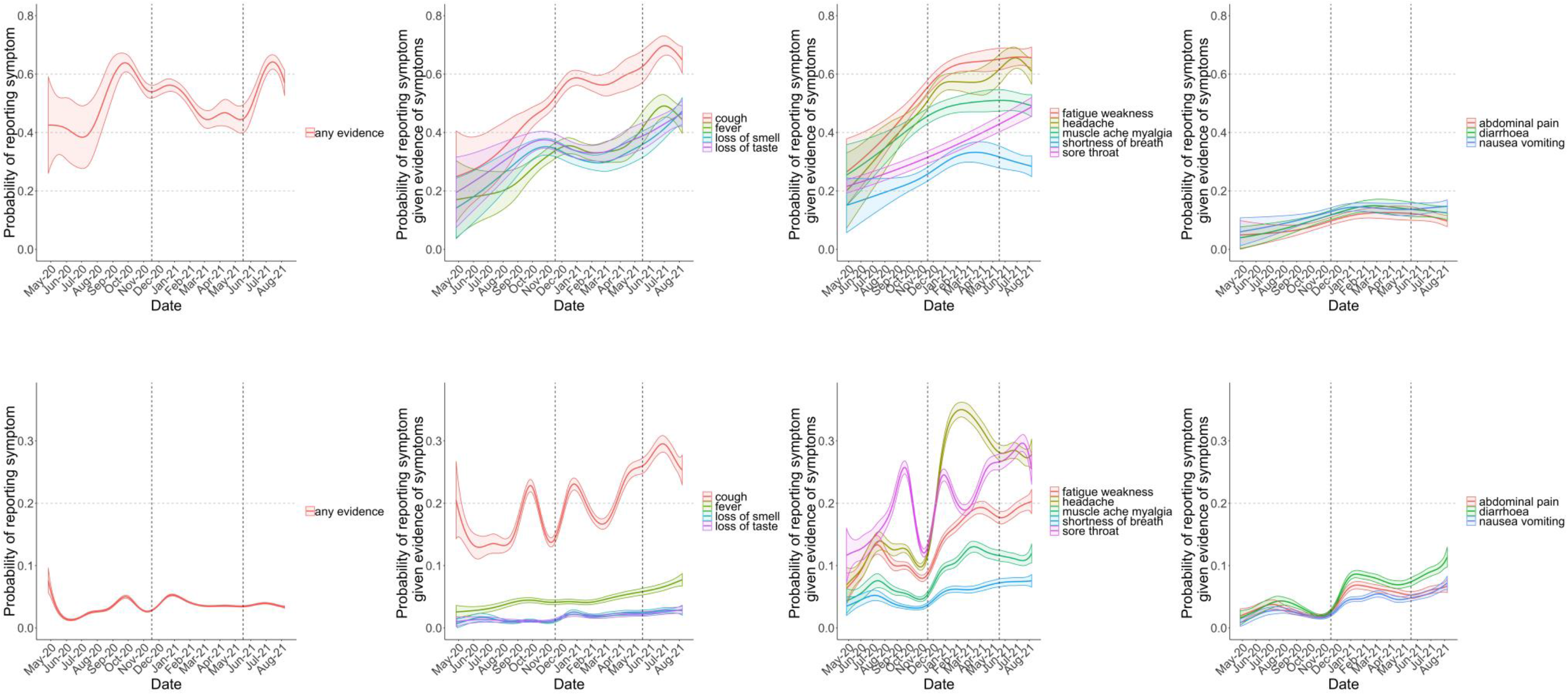
Probability of reporting any evidence of symptoms (first column), and specific classic symptoms (second column), gastrointestinal symptoms (fourth column) and other symptoms (third column) in those with evidence of symptoms, by calendar time in positive episodes (**top row**) and negative visits (**bottom row**). Note: models adjusted for age, sex, ethnicity (presented at the reference category age 45, male, white). The top and bottom rows have different scales for the y-axis. Dashed lines at 17 November 2020 and 17 May 2021 indicate the emergence of Alpha and Delta respectively, see **Fig.S1**.

### Symptomatology by age, sex and ethnicity

All symptoms showed marked variation across age in both positive episodes and negative visits, mostly being reported less in children and elderly adults (**Fig.3**). Loss of taste/smell were most frequently reported in symptomatic positive episodes in those aged ∼20y, decreasing gradually at older ages; both were rarely reported amongst symptomatic negative visits, consistent with their high specificity for SARS-CoV-2, although slightly more in the elderly. Sore throat and headache also peaked in symptomatic positive episodes and negative visits in late adolescence, but were common regardless of positivity. High proportions reported cough, relatively similarly amongst symptomatic positive episodes and negative visits to ∼10y, therefore without discriminating; however, above 20y the proportion reporting cough was more than double in symptomatic positive episodes, and increased to ∼60y, as did fatigue/weakness, shortness of breath, and diarrhoea.

**Figure 3.**
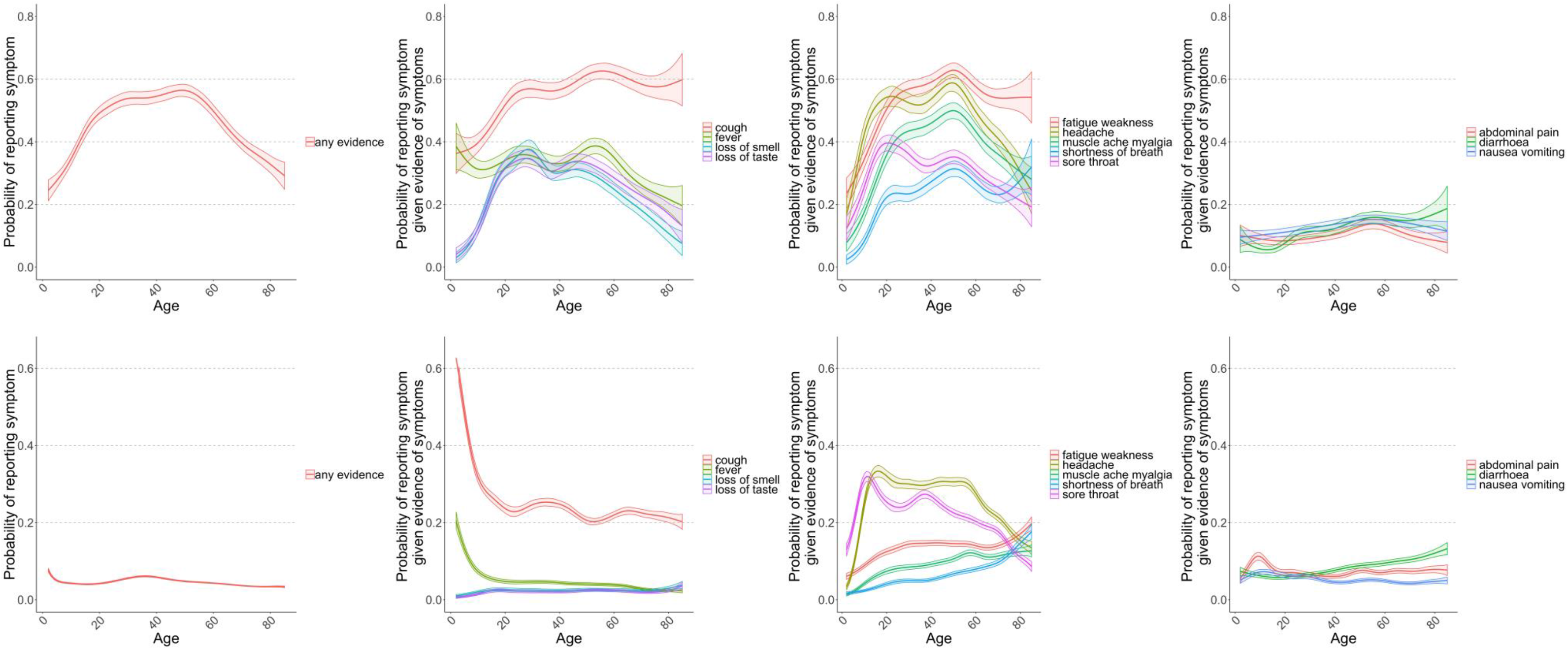
Probability of reporting any evidence of symptoms (first column), and specific classic symptoms (second column), gastrointestinal symptoms (fourth column) and other symptoms (third column) in those with evidence of symptoms, by age in positive episodes (**top row**) and negative visits (**bottom row**). Note: models adjusted for calendar date, sex, ethnicity (reference category 1 January 2021, male, white). The top and bottom row have different scales for the y-axis.

Adjusting for calendar time, age and ethnicity, women were generally more likely than men to report most symptoms (**Fig.4**). Increased reporting in women was significantly greater in symptomatic positive episodes than negative visits for loss of smell and taste, diarrhoea and shortness of breath, and significantly smaller for headache and sore throat (all heterogeneity p<0.01). Whilst there was no evidence of differential reporting of fever between male and female symptomatic negative visits, female symptomatic positive episodes were significantly less likely to report fever (p<0.001).

**Figure 4.**
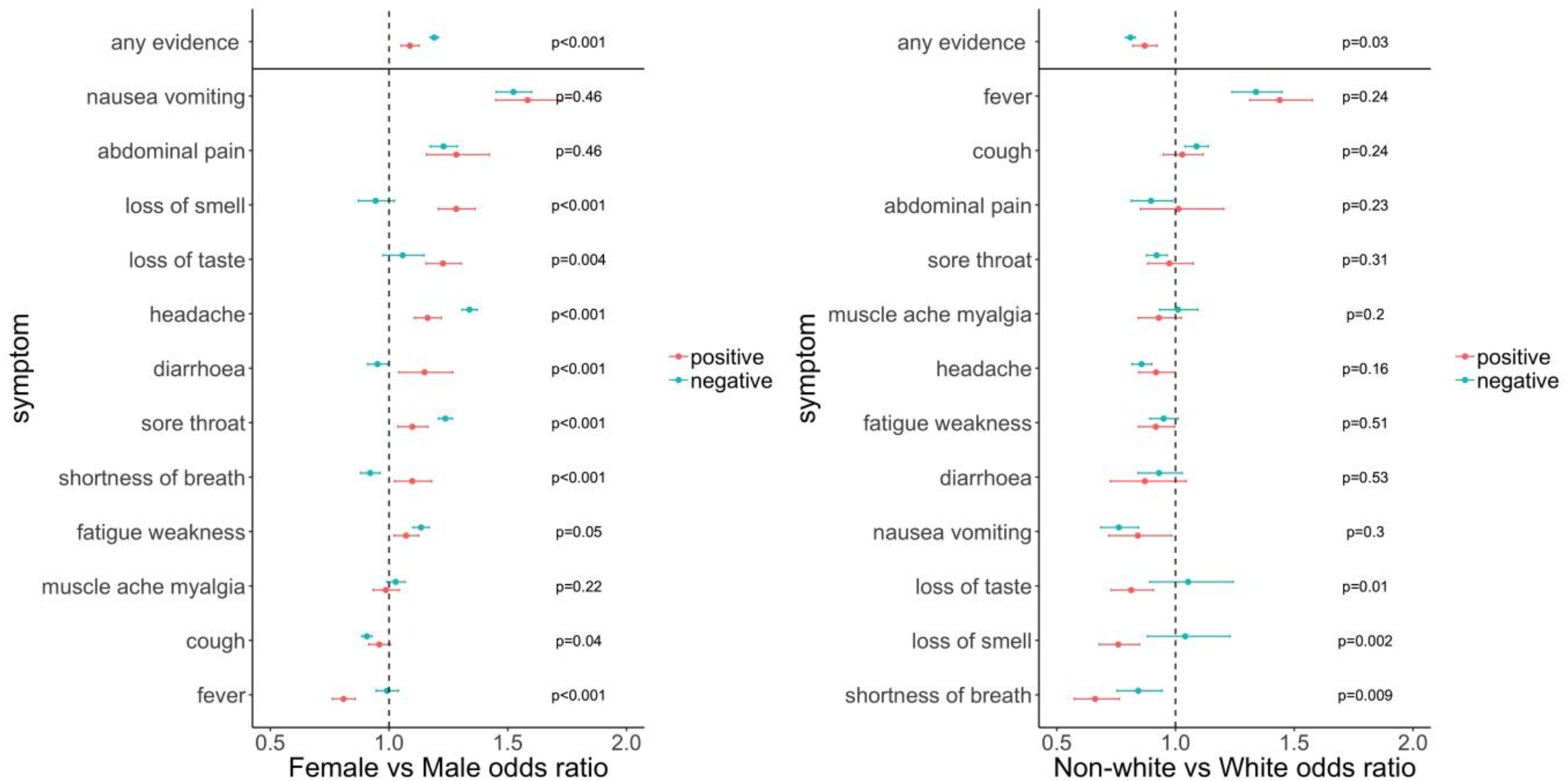
Odds ratios (95% CI) of reporting any evidence of symptoms, as well as each of the 12 symptoms in those with evidence of symptoms, in positive episodes (red) and negative visits (turquoise) by sex (female vs male, **left**), and ethnicity (non-white vs white, **right**). p-values are heterogeneity tests between the effects of sex and ethnicity on reporting symptoms in positive episodes vs negative visits. Note: models adjusted for calendar date (Fig.2), age (Fig.3), sex and ethnicity. Where 95% CI cross 1, there is no evidence that sex/ethnicity affects the odds of reporting that symptom **given evidence of symptoms** in positive episodes/negative visits. Where there is evidence of heterogeneity, there is a different effect of sex/ethnicity on reporting the symptom in positive episodes vs negative visits.

After adjusting for calendar time, age and sex, in both symptomatic positive episodes and negative visits, non-white ethnic groups were more likely to report fever than white ethnic groups, and less likely to report headache, nausea/vomiting and shortness of breath (**Fig.4**). Those from non-white ethnic groups were less likely to report loss of taste or smell and shortness of breath to a significantly greater degree in symptomatic positive episodes versus negative visits (all p<0.01).

### Symptoms by vaccination status

Both positive episodes and negative visits had lower odds of reporting any evidence of symptoms compared to those pre-vaccination with no evidence of difference (p>0.44, **Fig.5**). However, positives episodes ≥21d from first vaccination had significantly lower odds of reporting 10/12 symptoms compared to negative-visits (p<0.01).

**Figure 5.**
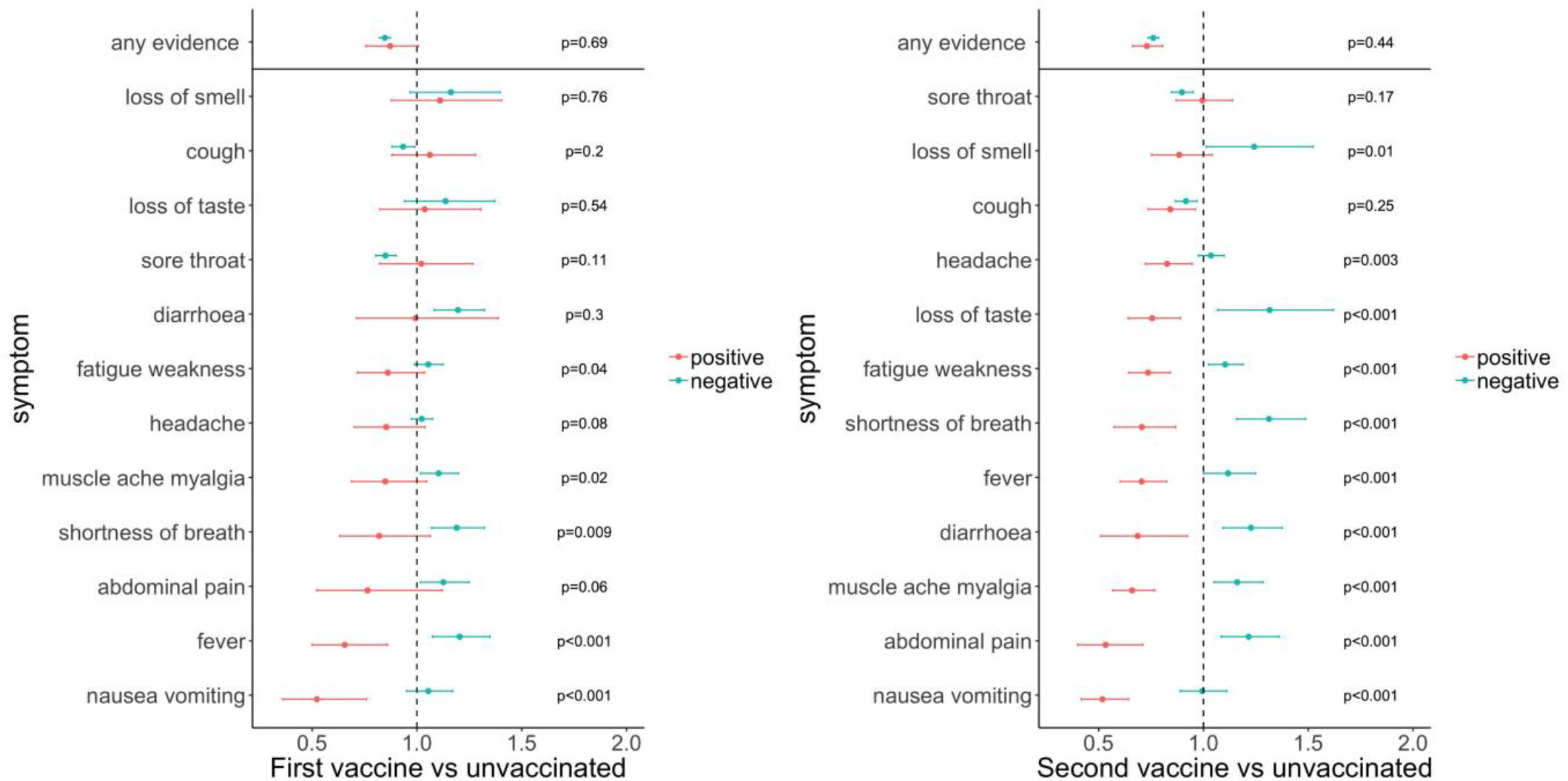
Odds ratios (95% CI) of reporting any evidence of symptoms, as well as each of the 12 symptoms in those with evidence of symptoms, in positive episodes (red) and negative visits (turquoise) by vaccination status (episodes/visits ≥21 days from 1^st^ vaccine and before 2^nd^ vaccine vs pre-vaccination, **left**, and ≥14 days from 2^nd^ vs pre-vaccination, **right**). p-values are heterogeneity tests between the effects of vaccination on reporting symptoms in positive episodes vs negative visits. Note: models adjusted for calendar date (Fig.2), age (Fig.3), sex (Fig.4) and ethnicity (Fig.5). Where 95% CI cross 1, there is no evidence that sex/ethnicity affects the odds of reporting that symptom **given evidence of symptoms** in positive episodes/negative visits. Where there is evidence of heterogeneity, there is a different effect of sex/ethnicity on reporting the symptom in positive episodes vs negative visits.

### Symptoms by Ct value in PCR-positive episodes

At low Ct values (≤20) (a proxy for high viral load), the most commonly reported symptoms were cough, fatigue/weakness, headache and muscle ache/myalgia, all occurring in >50% of episodes (**Fig.6**; adjusted for gene positivity pattern in **Fig.S3**). Above Ct>27.5 (reflecting lower viral loads), all symptoms declined in prevalence with a trajectory that tracked Ct; between 20-27.5, most symptoms showed little variation. Interestingly, prevalence of reported loss of taste/smell increased substantially from ∼30% to ∼45% between Ct 15-27.5, with smaller increases for shortness of breath.

**Figure 6.**
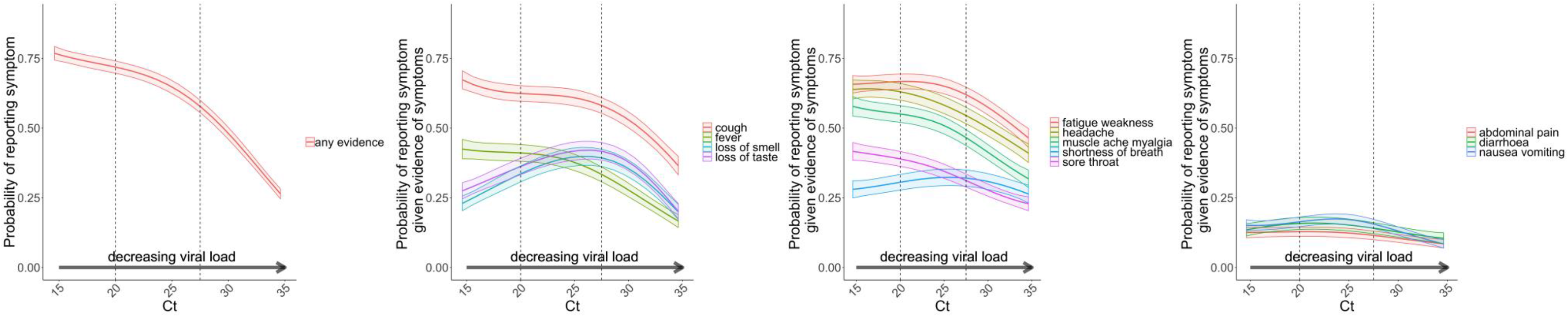
Probability of reporting any evidence of symptoms (first column), and the probability of reporting each of the 12 symptoms in those with evidence of symptoms, by Ct value Note: models adjusted for calendar date (Fig.2), age (Fig.3), sex and ethnicity (Fig.4) (reference category 1 January 2021, 45, male, white). See **Fig.S3** for models also adjusting for S-gene positivity pattern with similar results

### Symptom combinations predicting symptomatic PCR-positive episodes

Over the whole study, PPV of any evidence of symptoms for identifying PCR-positive episodes was 9%. Including any of the 12 elicited symptoms by definition maximised sensitivity (90% of symptomatic positive episodes reported at least one specific symptom; remainder reported any other symptom considered COVID-19 related only), but substantially increased TPC (8.7) and number of tests (2.3-fold) compared with using the four classic (cough, fever, loss of taste/smell) symptoms (74%, 4.6, 1-fold). For a fixed number of 1-8 symptoms, the choice of whether to maximise sensitivity or area under the receiver operating characteristic (AUROC) curve, or to minimise TPC or the inflation factor, led to different optimal combinations (**Table S4**). However, these frequently included one or more of the classic four symptoms. Sensitivity was generally higher for combinations including fatigue/weakness and/or headache, but at a cost of higher TPC, particularly for headache. Including gastrointestinal symptoms had the lowest TPC and numbers to be tested, but lowest sensitivity. In those ≥14 days post second dose, sore throat had similar effects on sensitivity and TPC as headache, and in children diarrhoea had a greater benefit for sensitivity (**Table S4**).

Balancing different performance metrics, adding fatigue/weakness to the classic four symptoms improved sensitivity from 74% to 81%, while dropping the AUROC by <0.01 (0.734 to 0.727) (**Fig.7**). However, TPC increased from 4.6 to 5.3, and 1.3 times more people would need testing. This combination generally performed well across subgroups (**Table S5**). Adding other symptoms to the classic four symptoms generally led to lower AUROCs, and at best similar sensitivity (**Table S5**, **Fig.7**), excepting children/adolescents in whom adding headache achieved highest sensitivities when considering adding only one extra symptom, and also highest AUROC for those aged under 10y (**Fig.S4-S8**).

**Figure 7.**
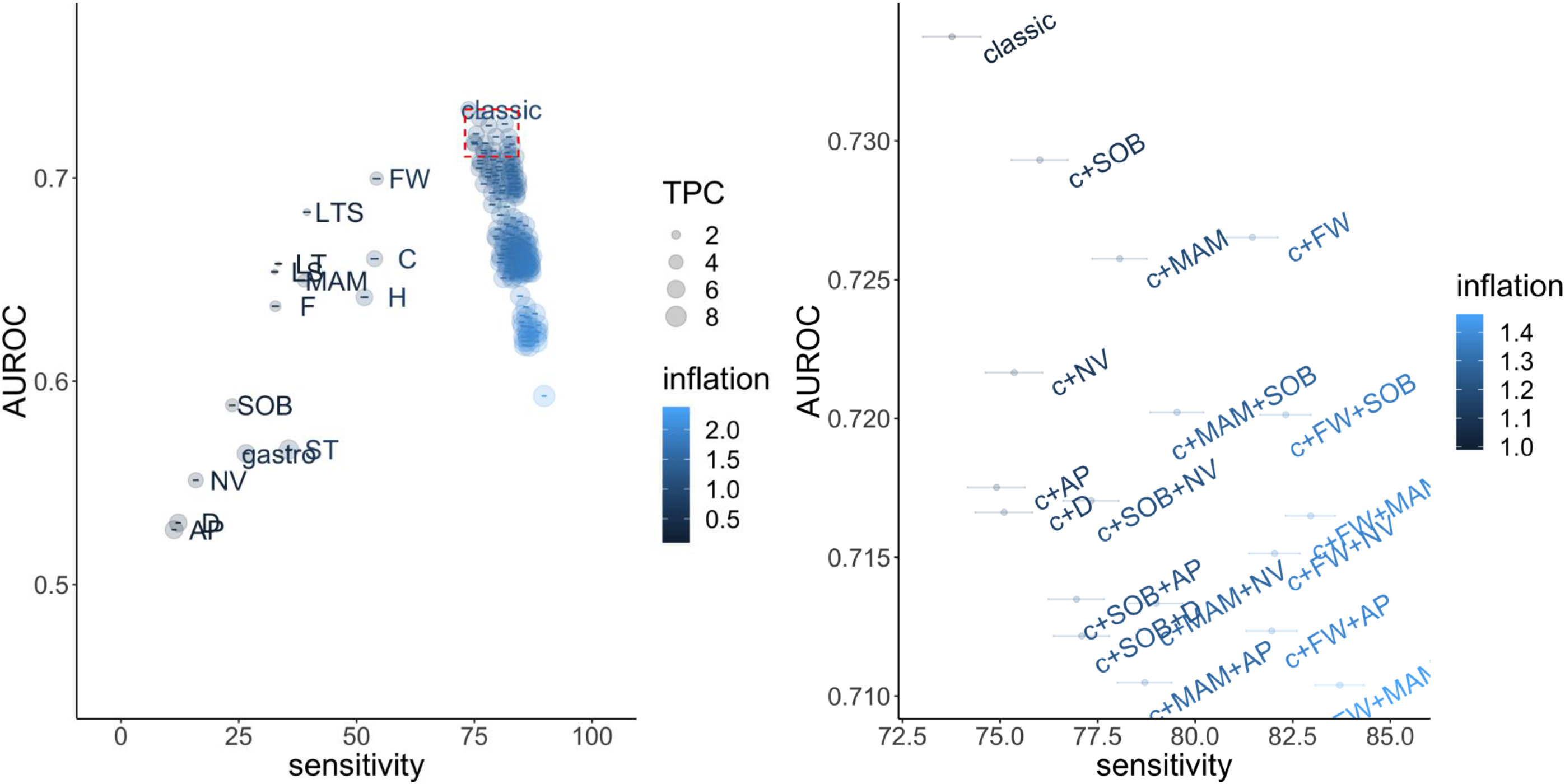
Performance of individual symptoms, as well as the classic four symptoms (cough, fever, loss of taste/smell), classic plus all possible combinations of 1/2/3/4 symptoms, and any of the 12 named symptoms, in predicting SARS-CoV-2 positivity in those with evidence of symptoms in terms of sensitivity and overall accuracy (AUROC). Note: Right hand panel is an expanded version of the top right corner of the left panel (red box, AUROC >90^th^ quantile, sensitivity > sensitivity of combination of classic 4 symptoms). Inflation (relative numbers reporting these symptoms compared to classic symptoms) and tests per positive case (TPC) are also included in the visualisation. TPC=1/positive predictive value. By definition, as the number of symptoms increases, sensitivity also increases. Note: abbreviations: c – classic, Fever - F, Headache - H, Muscle ache/myalgia - MAM, Weakness/tiredness - FW, Nausea/vomiting - NV, Abdominal pain - AP, Diarrhoea - D, Sore throat - ST, Cough - C, Shortness of breath - SOB, Loss of taste - LT, Loss of smell – LS, Loss of taste or smell – LTS

### Symptomatology at different stages of SARS-CoV-2 infection

Considering symptoms over time in positive episodes with ≥2 visits within 35 days (**Table S6**), the most common symptoms presenting after the index positive were fatigue/weakness (8%), headache (7%), cough (6%), loss of taste (6%), loss of smell (5%) or muscle ache/myalgia (5%) (**Table S7**). For most symptoms, Ct values were highest in those never reporting the symptom, lowest in those reporting it initially and subsequently, and intermediate where symptoms were reported at either the initial or subsequent visits only (**Fig.S9**). The main contrast was loss of taste and loss of smell, where Ct values were lowest in those reporting loss of taste or smell at subsequent visits only (p<0.0001).

## DISCUSSION

In a large, randomly selected community-based survey, we demonstrate that overall reporting of any symptoms in new SARS-CoV-2 positives varied substantially over calendar time (40-70%), reflecting changing dominance of specific variants (Alpha, Delta) and positivity rates (higher viral burden (low Ct)/symptomatic infections being identified more frequently at (mostly) monthly visits when rates are increasing[5]), but also background incidental changes (e.g. public awareness of SARS-CoV-2-associated symptoms, seasonal pathogens, schools re-opening). Additional important differences in the relative frequency of symptom reporting in positive episodes vs negative visits were also observed by age, sex, ethnicity and vaccination status. Broadly however, of the 12 symptoms evaluated, the four classic symptoms (fever, cough, loss/change of smell/taste) gave close to optimal symptom-based screening, for testing given limited capacity; where additional testing capacity is available adding fatigue/weakness had the greatest improvement in sensitivity (7%) whilst inflating TPC by only 15%.

Whilst the CDC approach of using a broad range of symptoms to prompt testing maximises sensitivity, this approach is associated with substantially higher TPC (8.7 vs 4.6 for classic symptoms) and total tests needed (2.3-fold) with associated costs and capacity requirements. The UK approach of focussing on four classic symptoms has lower sensitivity (74% vs 90%), but higher accuracy to detect SARS-CoV-2 symptomatic infection overall (AUROC 0.734 vs 0.593). Increases in sensitivity from adding symptoms to the four classic symptoms typically either reduced overall accuracy and/or increased TPC and tests needed, highlighting the importance of evaluating several test metrics. Although advantages from including any additional symptoms were limited, those that best improved sensitivity across multiple subgroups included either fatigue/weakness or muscle ache/myalgia, or, in children/adolescents, headache. The REACT study[7] evaluating symptom constellations during the Alpha wave (December-2020/January-2021) suggested adding headache, muscle aches, chills and appetite loss to the classic symptoms. Our survey did not specifically elicit chills or appetite loss, which may partially overlap with fever and nausea/vomiting/abdominal pain; however, we found headache to have poorer specificity (i.e. to be commonly reported in test-negatives), particularly in adults, leading to substantially increased TPC despite improving sensitivity. REACT also suggested having different symptom sets for adults and children in order to optimise sensitivity, which would require careful public health messaging. ZOE[8] suggested an algorithm also including working in healthcare; whilst this could theoretically be programmed into an online test system, such complexity risks gaming the system if individuals cannot otherwise get a test.

The main limitation is that the survey collected information on only 12 specific COVID-19 symptoms, plus one generic question, to minimise participant burden. We therefore could not evaluate some symptoms more recently proposed for inclusion in expanded case definitions, such as coryza[13,14]. Parents/carers reported symptoms for children; symptom reporting may be affected by other cultural differences we could not adjust for as well as public awareness (e.g. increased reporting of loss of taste/smell once this became a “recognised” symptom). Power was limited within some subgroups, e.g. children and specific non-white ethnic groups. Our survey does not include those in care homes and with severe disease admitted to hospital who may have different symptom profiles. Testing was predominantly monthly; although individuals were followed longitudinally, we had limited resolution to assess the short-term evolution of symptoms during an infection.

The main study strengths are its size and population representativeness, particularly capturing episodes of mild infection in the community. We took a stringent approach to defining our ‘test-negative’ comparator to limit possible contamination from undetected positives/ongoing COVID-19. We report over periods that include different dominant viral variants. We took a pragmatic approach to comparing test performance, taking into account trade-offs between overall accuracy, sensitivity and TPC over different background prevalences, reflecting practical concerns regarding testing capacity, rather than optimising individual criteria.

Overall, we did not find any major shift away from the importance of the classic four symptoms in positive episodes with the emergence of the Delta variant and vaccine roll-out in the UK. Given their concurrent changes in test-negatives, recent reports of associations with sore throat may reflect background increases in other respiratory infections/hayfever, potentially even with SARS-CoV-2 isolated incidentally given that one-third of cases are estimated to be asymptomatic[2]. Currently, we therefore have limited evidence for expanding the case definition beyond the classic four symptoms where universal testing is not practical/affordable, with fatigue/weakness the most promising candidate. However, this requires ongoing monitoring as other respiratory viruses increasingly circulate following lifting of restrictions with vaccine roll-out[15–18], potentially altering the specificity of symptoms in determining SARS-CoV-2 vs other community-acquired infections.

## Data Availability

Data are still being collected for the COVID-19 Infection Survey. De-identified study data are available for access by accredited researchers in the ONS Secure Research Service (SRS) for accredited research purposes under part 5, chapter 5 of the Digital Economy Act 2017. For further information about accreditation, contact or visit the SRS website.

## Contributors

This specific analysis was designed by ASW, KBP, PCM, NS, DWE, TH, DC, TEAP, K-DV. K-DV conducted the statistical analysis of the survey data. K-DV, NS, PCM, ASW drafted the manuscript. All authors contributed to interpretation of the study results, and revised and approved the manuscript for intellectual content. K-DV is the guarantor and accepts full responsibility for the work and conduct of the study, had access to the data, and controlled the decision to publish. The corresponding author (K-DV) attests that all listed authors meet authorship criteria and that no others meeting the criteria have been omitted.

## Ethical approval

The study received ethical approval from the South Central Berkshire B Research Ethics Committee (20/SC/0195).

## Transparency

The lead authors affirm that the manuscript is an honest, accurate, and transparent account of the study design being reported, no important aspects of the study have been omitted, and any discrepancies from the study as originally planned (and, if relevant, registered) have been explained. Dissemination to participants and related patient and public communities: Results of individual tests were communicated to the participants. Overall study results were disseminated through the preprint of the study. Findings were disseminated in lay language in the national and local press.

## Acknowledgements

We are grateful for the support of all COVID-19 Infection Survey participants. Office for National Statistics: Sir Ian Diamond, Emma Rourke, Ruth Studley, Tina Thomas. Office for National Statistics COVID-19 Infection Survey Analysis and Operations teams, in particular: Daniel Ayoubkhani, Russell Black, Antonio Felton, Megan Crees, Joel Jones, Lina Lloyd, Esther Sunderland. University of Oxford, Nuffield Department of Medicine: Ann Sarah Walker, Derrick Crook, Philippa C Matthews, Tim Peto, Emma Pritchard, Nicole Stoesser, Karina-Doris Vihta, Jia Wei, Alison Howarth, George Doherty, James Kavanagh, Kevin K Chau, Sarah Cameron, Phoebe Tamblin-Hopper, Magda Wolna, Rachael Brown, Stephanie B Hatch, Daniel Ebner, Lucas Martins Ferreira, Thomas Christott, Brian D Marsden, Wanwisa Dejnirattisai, Juthathip Mongkolsapaya, Sarah Hoosdally, Richard Cornall, David I Stuart, E Yvonne Jones, Gavin Screaton. University of Oxford, Nuffield Department of Population Health: Koen Pouwels. University of Oxford, Big Data Institute: David W Eyre, Katrina Lythgoe, David Bonsall, Tanya Golubchik, Helen Fryer. University of Oxford, Radcliffe Department of Medicine: John Bell. University of Manchester: Thomas House. Public Health England: John Newton, Julie Robotham, Paul Birrell. IQVIA: Helena Jordan, Tim Sheppard, Graham Athey, Dan Moody, Leigh Curry, Pamela Brereton. Glasgow Lighthouse Laboratory: Jodie Hay, Harper VanSteenhouse. National Biocentre: Anna Godsmark, George Morris, Bobby Mallick, Phil Eeles. Oxford University Hospitals NHS Foundation Trust: Stuart Cox, Kevin Paddon, Tim James, Sarah Cameron, Phoebe Tamblin-Hopper, Magda Wolna, Rachael Brown. Department of Health: Jessica Lee.

## Supplementary Material

### Supplementary Methods

The presence of three SARS-CoV-2 genes (ORF1ab, N, S) was identified using real-time polymerase chain reaction (RT-PCR) with the TaqPath RT-PCR COVID-19 kit (Thermo Fisher Scientific, Waltham, MA, USA), analysed using UgenTec Fast Finder 3.300.5 (TaqMan 2019-nCoV assay kit V2 UK NHS ABI 7500 v2.1; UgenTec, Hasselt, Belgium).

#### Choice of negative visits in the comparator group

As a comparator group, we initially included all visits where PCR tests were negative, and then excluded visits where symptoms could plausibly be related to ongoing effects of COVID-19 or long COVID, where there was a high pre-test probability that the participant actually had a new COVID-19 infection that had not been detected in the survey, or where symptoms were likely driven by recent vaccination. Specifically, we excluded all negative visits (numbers in **Table S1**):

1. **<u>From -90 days before</u>** the first S-antibody positive blood test in the study prior to vaccination, where such antibody results are likely to represent previous undetected infection (these results were available only in a random subset of the population);
2. **<u>From -35 days before</u>** the first swab positive onwards from individuals who ever tested PCR positive in the study or positive on either PCR or LFD in the linked English testing programme (to avoid ongoing long COVID symptoms,^2^ and COVID-related symptoms occurring shortly before the positive test);
3. **<u>From -35 days before</u>** any self-reported positive swab test result onwards (for the same reason; reflecting the fact that individuals may have obtained tests elsewhere)
4. From a small number of individuals who reported either loss of taste or loss of smell at their first study visit and had no national testing programme result within [-21,+21] days (all before 1 July 2020), given the high specificity of this symptom for COVID-19 infection, the fact that it would have been impossible for these individuals to get an external test at the time and the potential for subsequent symptoms to represent long COVID;
5. Where participants reported self-isolating OR contact with **<u>definite</u>** positives in the preceding 28 days (since these individuals have much higher risk of SARS-CoV-2 infection which may not have been detected) and the **<u>previous and the next visit</u>** (because of higher risk of unidentified positivity, and because they may have been contact traced through the national training programme they may be more likely to report symptoms through recall bias, regardless of status);
6. Occurring within [-7,+14 days] of either first or second vaccination date^3^, to avoid the inclusion of common symptoms caused by vaccination in the test-negative comparator group and to reflect the possibility of small inaccuracies in reported date of vaccination for some participants.

Time windows were arbitrary but aligned with other analyses or windows for considering symptoms associated with positive episodes.

#### Subgroups

In order to assess the impact of various changes over the course of the epidemic, we considered symptoms overall in all positive episodes, and in specific subgroups, as follows:

a. S-gene present before 17/Nov/2020 (wild-type) vs S-gene absent from 17/Nov/2020 to 17/May/2020 (Alpha-compatible) vs S-gene present from 17/May/2020 onwards (Delta-compatible) (**Fig.S1**); all restricted to Ct<30 to increase the chance that the survey positive test was closer to the start of the infection
b. Ct<30 or ≥30 as a proxy for higher viral load (where symptoms may be more completely ascertained if the infection is identified close to onset) vs lower viral load
c. Up to 0 days before 1^st^ vaccination date or unvaccinated, from 21 days post 1^st^ vaccination date to 13 days post 2^nd^ vaccination date inclusive, from 14 days post 2nd vaccination date onwards. Episodes/tests 0-20 days after first vaccination were excluded as symptoms may be due to side-effects.
d. Age groups 2-5, 6-10, 11-15, 16-44, 45-64, 65+ years
e. All positive episodes split between before 1/Sep/2020, 1/Sep/2020-17/Nov/2020, 17/Nov/2020-1/Mar/2021, 1/Mar/2021-17/May/2021, 17/May/2021-17/Jul/2021 on the basis of background incidental symptoms in negative visits (different background rates could be driven either by epidemic dynamics and/or other infections being more prevalent during certain periods)

#### Cycle threshold (Ct) values

Each positive test has a Ct value for each positive gene, leading to 1-3 individual Ct values per positive result. As the Spearman correlation between Ct values for each pair of genes (when present together) was very high (>0.98), we first took the arithmetic mean of all Ct values for detected genes for each positive as the single Ct value per positive test. We then took the minimum of these Ct values across all positive tests in an episode as the Ct value for each positive episode.

#### Generalised additive models

In regression models for reporting any evidence of symptoms and specific symptoms in those with evidence of symptoms in positive episodes and negative visits, we truncated age at 85y and Ct at the 5^th^ and 95^th^ percentiles in order to avoid undue influence of outliers. Age was modelled as smoothing spline. Due to small numbers (**Table S3**) we were only able to investigate differences by self-reported ethnicity as white vs non-white.

bam(cbind(n_withsymptom, n_withoutsymptom) ∼
s(study_day, bs=“bs”, k=15, by=Sars_COV_2_positivity) +
s(age_at_visit, bs=“bs”, k=15, by=Sars_COV_2_positivity) +
sex:Sars_COV_2_positivity + ethnicity_wo:Sars_COV_2_positivity + sex + ethnicity_wo +
Sars_COV_2_positivity, family=binomial(link=“cloglog”), method = “fREML”, data = data,
discrete=TRUE, nthreads =12)

bam(cbind(n_withsymptom, n_withoutsymptom) ∼
vaccinated:Sars_COV_2_positivity + vaccinated +
s(study_day, bs=“bs”, k=15, by=Sars_COV_2_positivity) +
s(age_at_visit, bs=“bs”, k=15, by=Sars_COV_2_positivity) +
sex:Sars_COV_2_positivity + ethnicity_wo:Sars_COV_2_positivity +
sex + ethnicity_wo + Sars_COV_2_positivity, family=binomial(link=“cloglog”), method =
“fREML”, data = data, discrete=TRUE, nthreads =12)

In positive episodes only:

bam(cbind(n_withsymptom, n_withoutsymptom) ∼
s(ct _mean, bs=“bs”, k=5) +
s(study_day, bs=“bs”, k=15) +
s(age_at_visit, bs=“bs”, k=15) +
sex + ethnicity_wo, family=binomial(link=“cloglog”), method = “fREML”, data = data,
discrete=TRUE, nthreads =12)

Also adjusting for S-gene positivity (present/absent/unknown):

bam(cbind(n_withsymptom, n_withoutsymptom) ∼
s(ct _mean, bs=”bs”, k=5) +
S_gene_positivity_pattern +
s(study_day, bs=”bs”, k=15) +
s(age_at_visit, bs=”bs”, k=15) +
sex + ethnicity_wo, family=binomial(link=”cloglog”), method = “fREML”, data = data,
discrete=TRUE, nthreads =12)

#### Performance metrics

For each combination of symptoms considered, we calculated the number of true positives (≥1 symptom from the combination being considered reported for a positive episode; TP), false positives (≥1 symptom reported for a negative visit; FP), true negatives (symptoms in the combination not reported for a negative visit; TN), false negatives (symptoms in the combination not reported in a positive episode; FN). We compared sensitivity=TP/(TP+FN); specificity=TN/(TN+FP); positive predictive value=TP/(TP+FP) (PPV); negative predictive value=TN/(TN+FN) (NPV); area under the receiver operating characteristic curve (AUROC), the area under the curve of specificity and 1–sensitivity, which, in this case where we have a binary outcome and a binary exposure, is 0.5*(sensitivity+specificity); tests per case identified=1/PPV (TPC); and the inflation factor=episodes/visits with ≥1 symptom/episodes/visits with classic symptoms.

## Results

### Defining the negative visit comparator group

**Table S1** shows the total number of test-negative visits and the number of participants in which they occur, in all PCR negative visits and restricting to those visits with evidence of symptoms only, for each step of the hierarchical restriction of test-negative visits to form the comparator. This was done in order to exclude potential contamination from undetected positives, ongoing symptoms after detected positives, and symptoms due to vaccination. Overall, 75% of test-negative visits in 93% participants with any test-negative visit were retained in analyses, and 62% of test-negative visits where any symptoms were reported in 68% of participants with any test-negative visit where any symptoms were reported. Most test-negative visits were excluded because of self-isolation or contact with definite positives. The larger exclusion of test-negative visits where any symptoms were reported, despite the fact that symptoms were not a reason for exclusion (with the exception of visits from a small number of individuals reporting loss of taste/smell in 2020 before tests were available) illustrate the likely contamination of all test-negative visits with both undetected positives and ongoing symptoms post COVID-19 infection (**Table S2**).

Comparing the probability of reporting specific symptoms in negative visits with evidence of symptoms over time before (**Fig.S10**) and after (**Fig.2**) the final exclusion of negative visits that occurred within [-7,+14 days] of either first or second vaccination date, the substantially higher probabilities of reporting fever, headache, fatigue weakness, muscle ache myalgia and nausea-vomiting in the first quarter of 2021 are plausibly driven by side effects of vaccination, as these probabilities markedly reduce after the exclusion.

---

**Figure S1.**
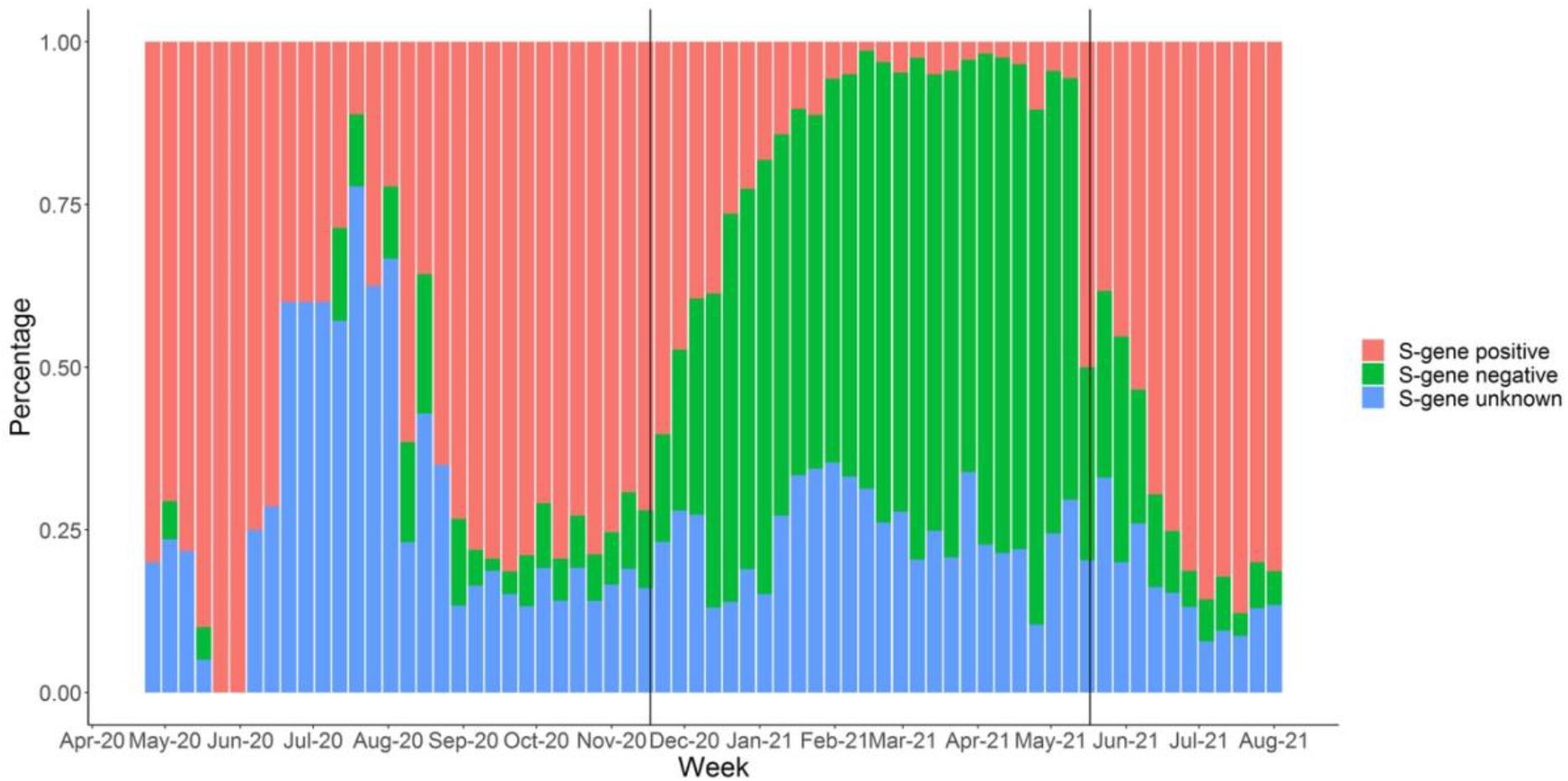
Percentage of positive episodes with each S-gene positivity pattern over time Note: vertical lines at 17 November 2020 and 17 May 2021. S-gene positive if the S-gene was ever detected within any positive test in the episode (by definition, with N/ORF1ab/both) (wild-type/Delta-compatible), otherwise S-gene negative if positive at least once for ORF1ab+N (Alpha-compatible), otherwise S-gene unknown (N-only/ORF1ab-only).

**Figure S2.**
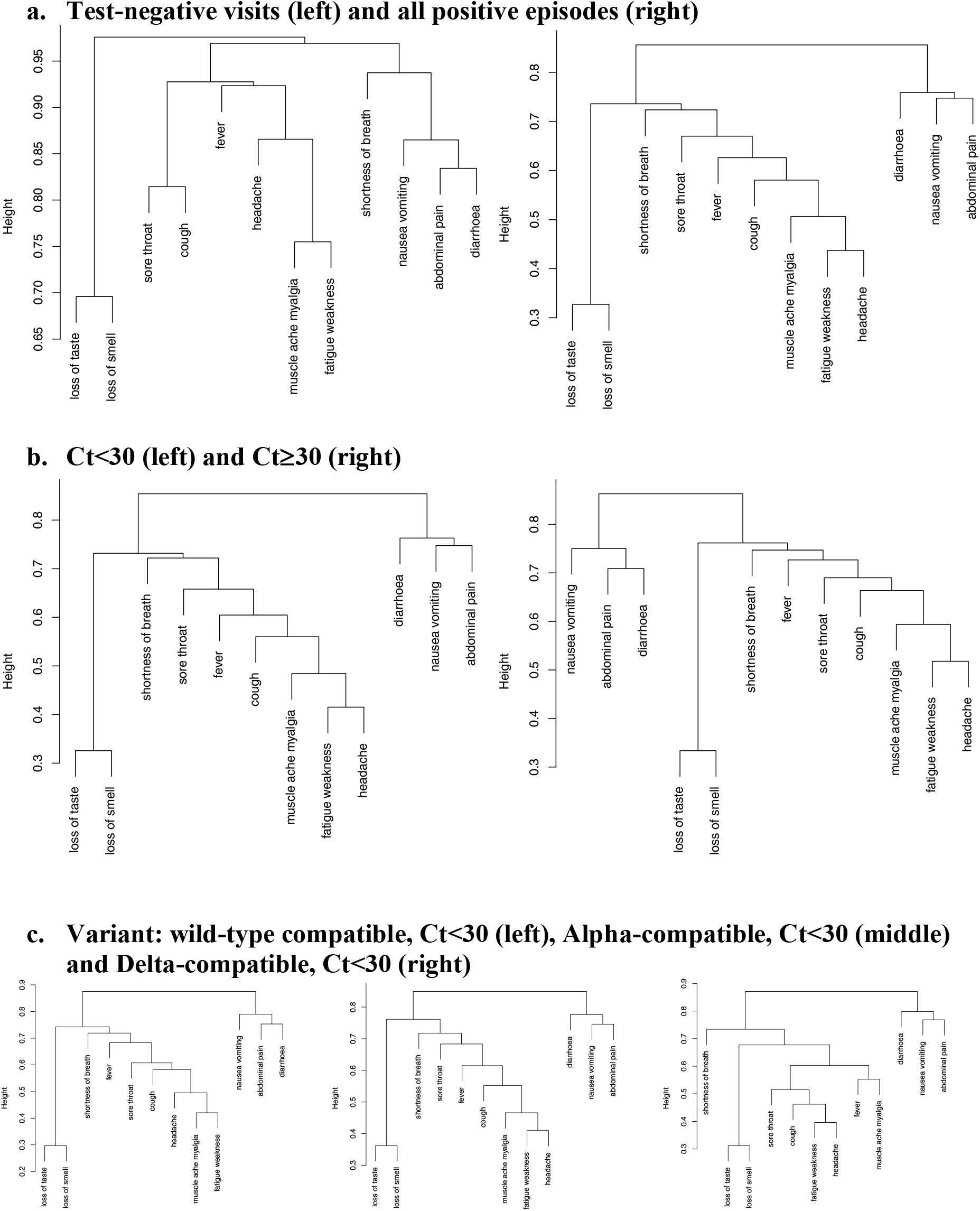
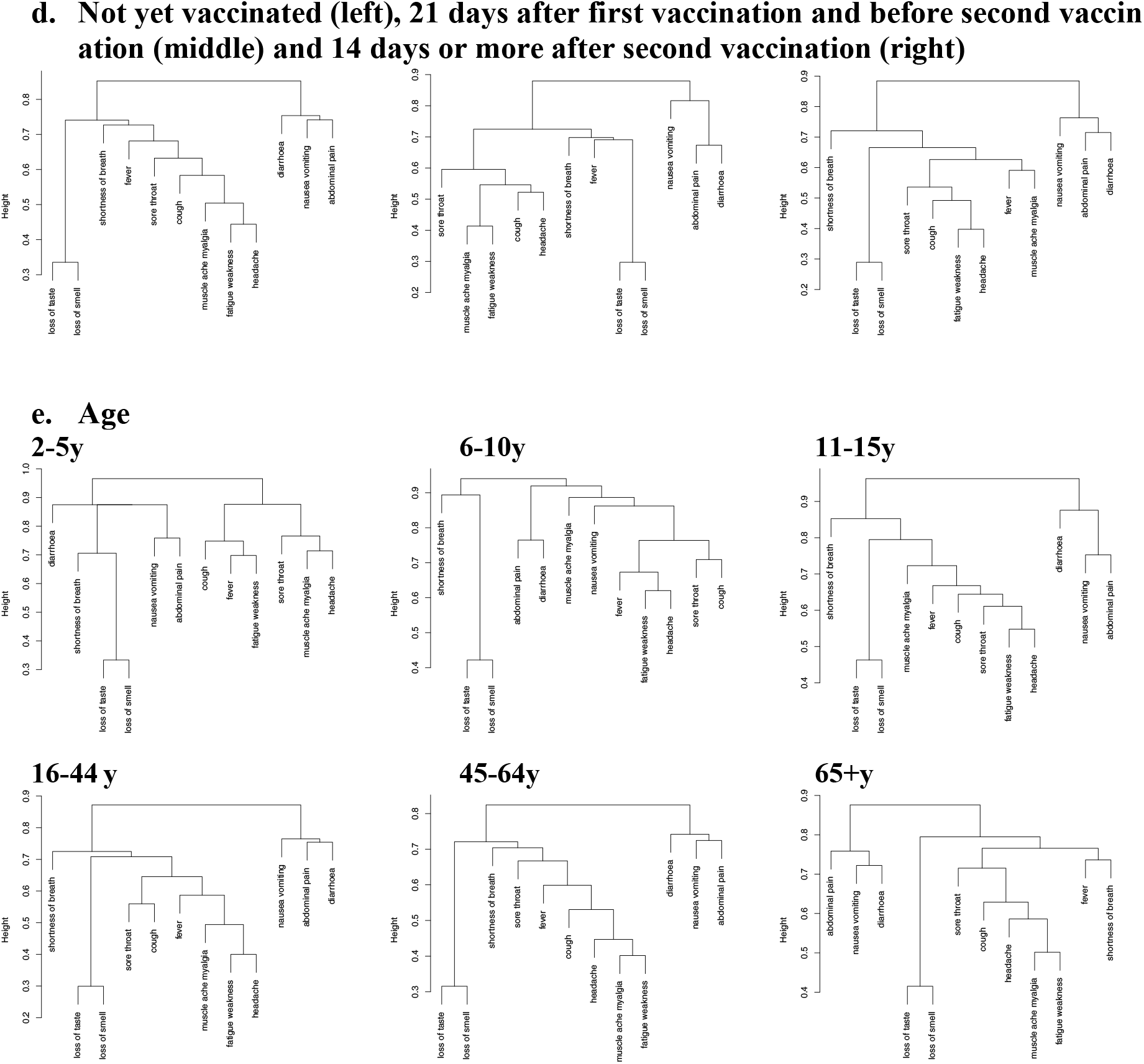
Hierarchical clustering of symptoms Note: different scales.

**Figure S3.**
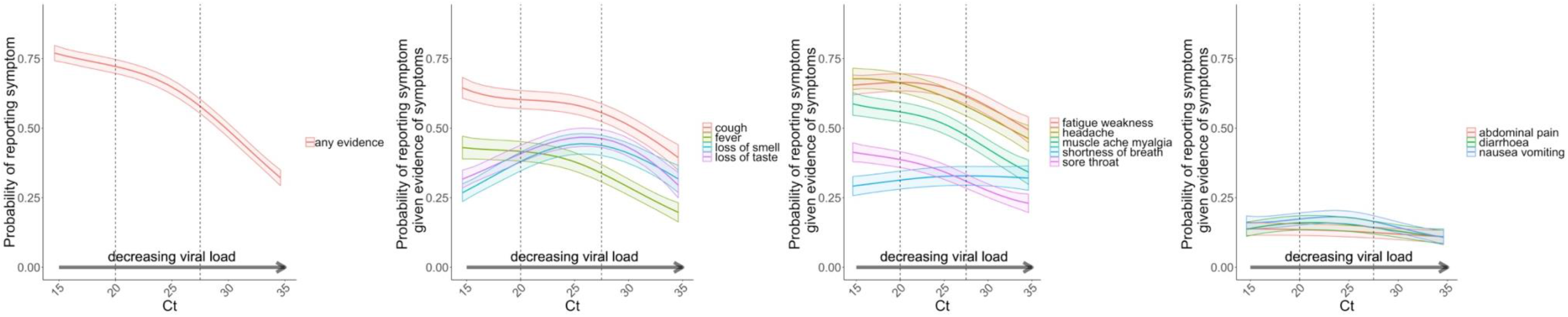
Probability of reporting any evidence of symptoms, as well as the probability of reporting each of the 12 symptoms in those with evidence of symptoms by mean Ct (model adjusted for age, sex and ethnicity and S-gene positivity pattern).

**Figure S4.**
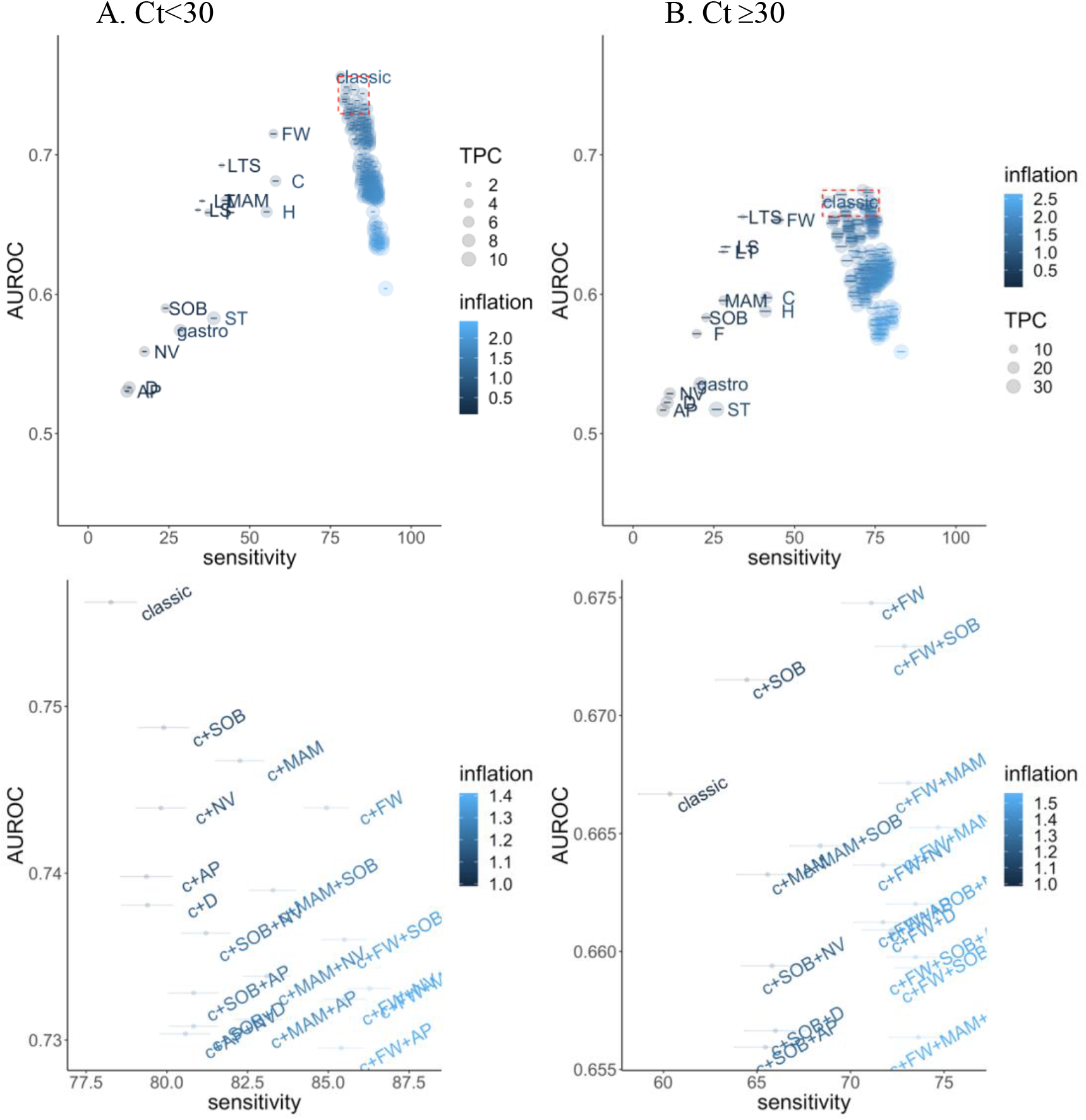
Performance of individual symptoms, as well as the classic four symptoms (cough, fever, loss of taste/smell), classic plus all possible combinations of 1/2/3/4 symptoms, and any of the 12 named symptoms, in predicting SARS-CoV-2 positivity in those with evidence of symptoms in terms of sensitivity and overall accuracy (AUROC). Note: Bottom row is an expanded version of the top right corner of the top row panels (red box, AUROC >90^th^ quantile, sensitivity > sensitivity of combination of classic 4 symptoms). Inflation (relative numbers reporting these symptoms compared to classic symptoms) and tests per positive case (TPC) are also included in the visualisation. TPC=1/positive predictive value. By definition, as the number of symptoms increases, sensitivity also increases. Note: abbreviations: c – classic, Fever - F, Headache - H, Muscle ache/myalgia - MAM, Weakness/tiredness - FW, Nausea/vomiting - NV, Abdominal pain - AP, Diarrhoea - D, Sore throat - ST, Cough - C, Shortness of breath - SOB, Loss of taste - LT, Loss of smell – LS, Loss of taste or smell – LTS

**Figure S5.**
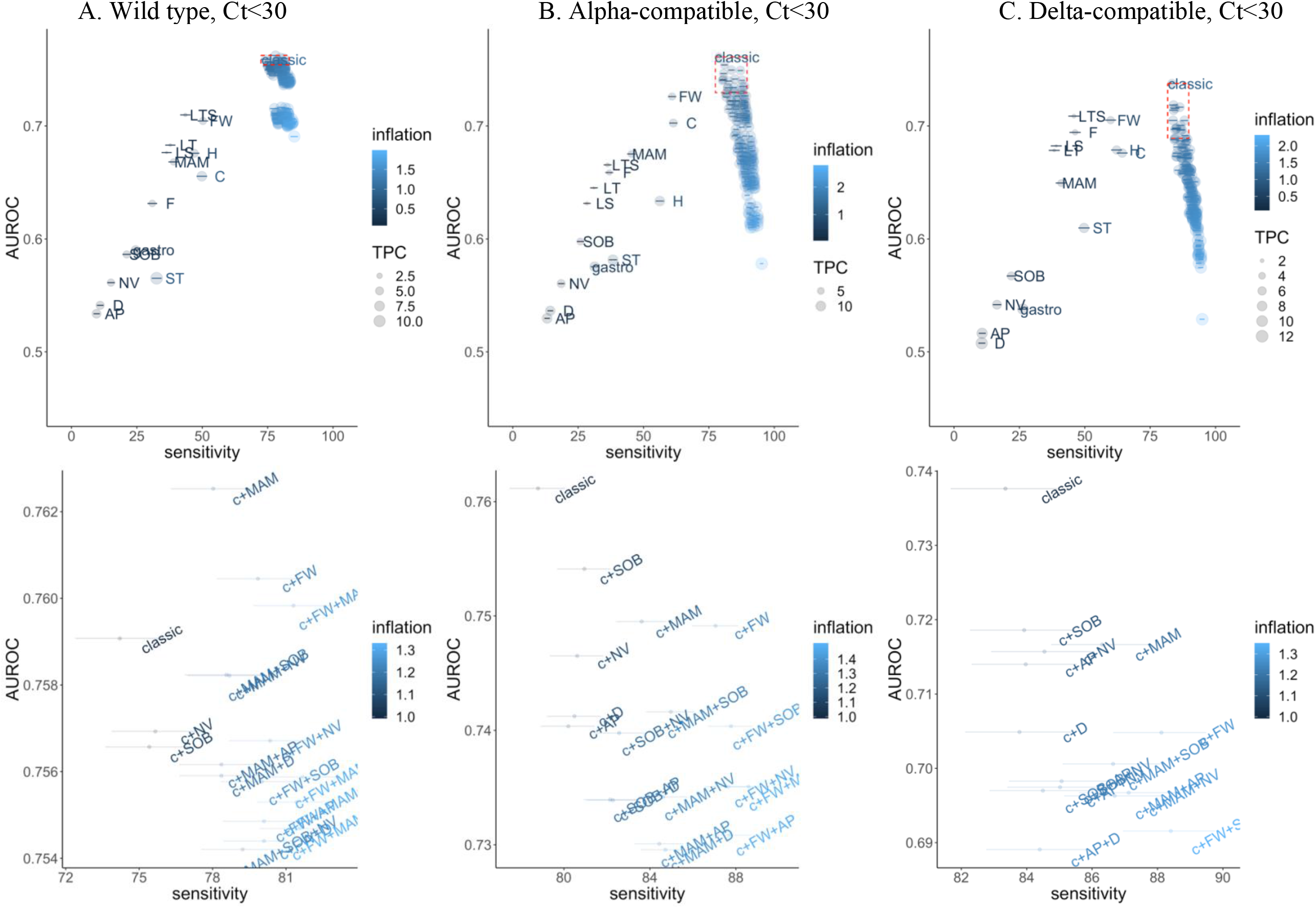
Performance of individual symptoms, as well as the classic four symptoms (cough, fever, loss of taste/smell), classic plus all possible combinations of 1/2/3/4 symptoms, and any of the 12 named symptoms, in predicting SARS-CoV-2 positivity in those with evidence of symptoms in terms of sensitivity and overall accuracy (AUROC). Note: Bottom row is an expanded version of the top right corner of the top row panels (red box, AUROC >90^th^ quantile, sensitivity > sensitivity of combination of classic 4 symptoms). Inflation (relative numbers reporting these symptoms compared to classic symptoms) and tests per positive case (TPC) are also included in the visualisation. TPC=1/positive predictive value. By definition, as the number of symptoms increases, sensitivity also increases. Note: abbreviations: c – classic, Fever - F, Headache - H, Muscle ache/myalgia - MAM, Weakness/tiredness - FW, Nausea/vomiting - NV, Abdominal pain - AP, Diarrhoea - D, Sore throat - ST, Cough - C, Shortness of breath - SOB, Loss of taste - LT, Loss of smell – LS, Loss of taste or smell – LTS

**Figure S6.**
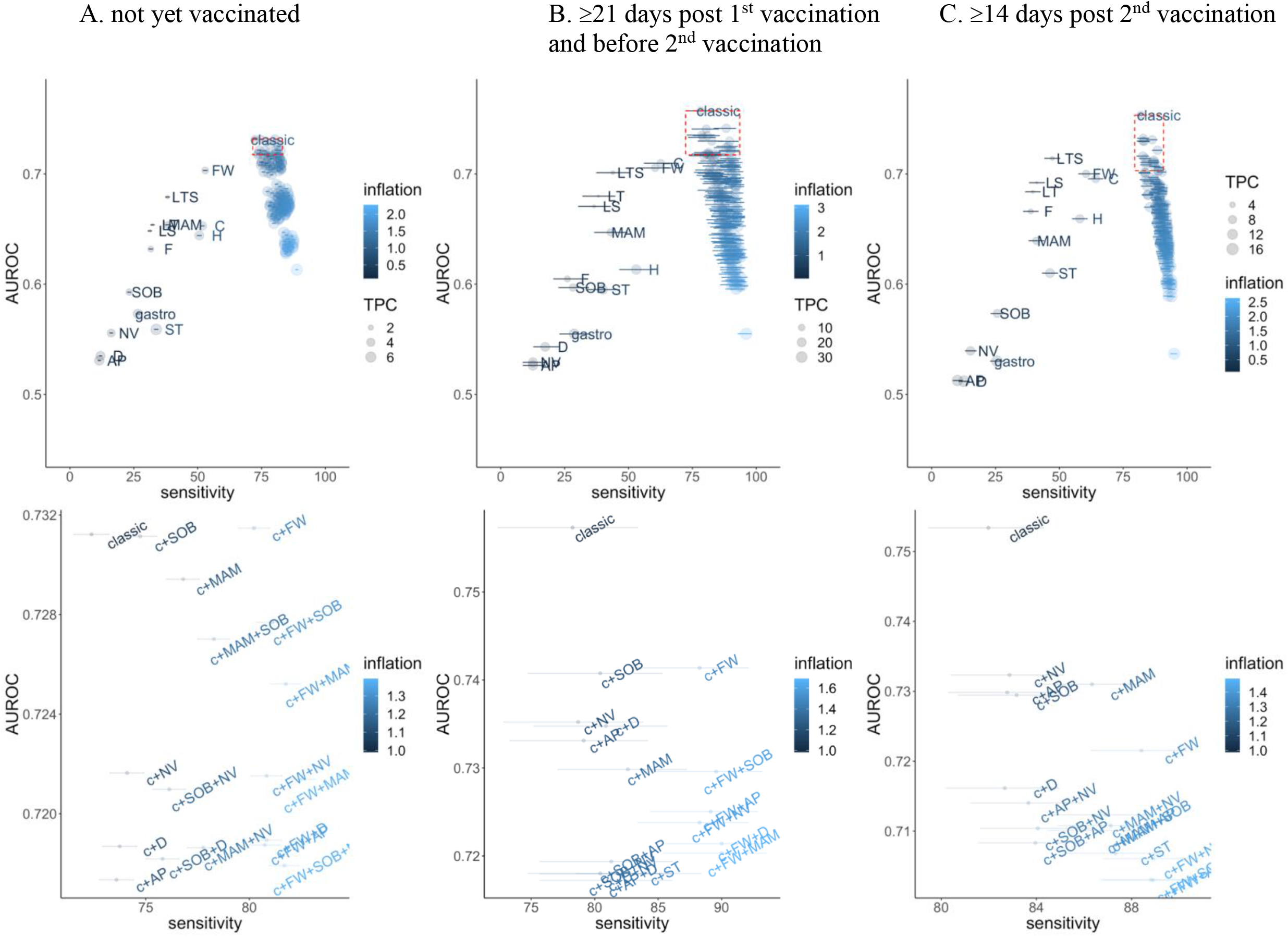
Performance of individual symptoms, as well as the classic four symptoms (cough, fever, loss of taste/smell), classic plus all possible combinations of 1/2/3/4 symptoms, and any of the 12 named symptoms, in predicting SARS-CoV-2 positivity in those with evidence of symptoms in terms of sensitivity and overall accuracy (AUROC). Note: Bottom row is an expanded version of the top right corner of the top row panels (red box, AUROC >90^th^ quantile, sensitivity > sensitivity of combination of classic 4 symptoms). Inflation (relative numbers reporting these symptoms compared to classic symptoms) and tests per positive case (TPC) are also included in the visualisation. TPC=1/positive predictive value. By definition, as the number of symptoms increases, sensitivity also increases. Note: abbreviations: c – classic, Fever - F, Headache - H, Muscle ache/myalgia - MAM, Weakness/tiredness - FW, Nausea/vomiting - NV, Abdominal pain - AP, Diarrhoea - D, Sore throat - ST, Cough - C, Shortness of breath - SOB, Loss of taste - LT, Loss of smell – LS, Loss of taste or smell – LTS

**Figure S7.**
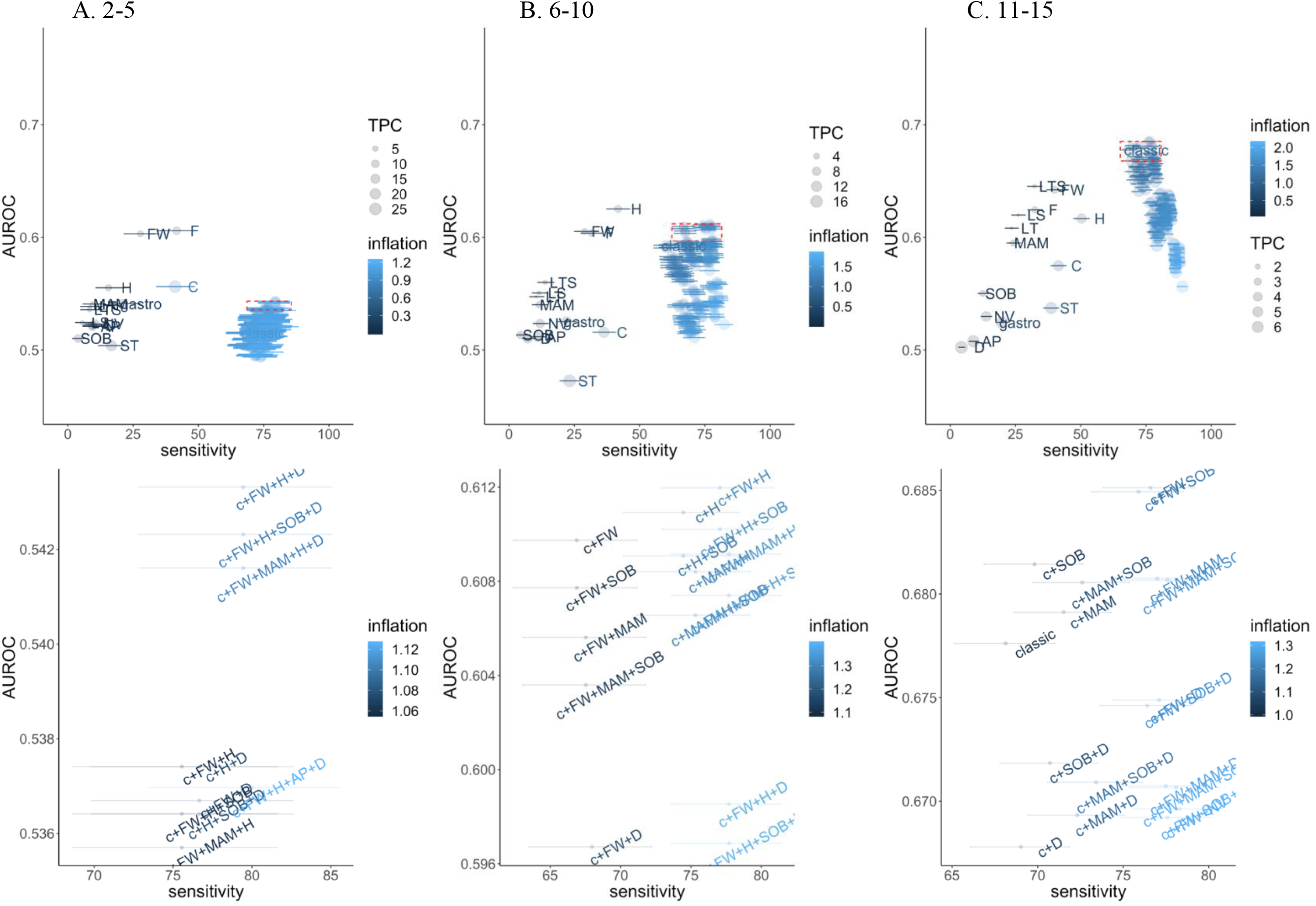
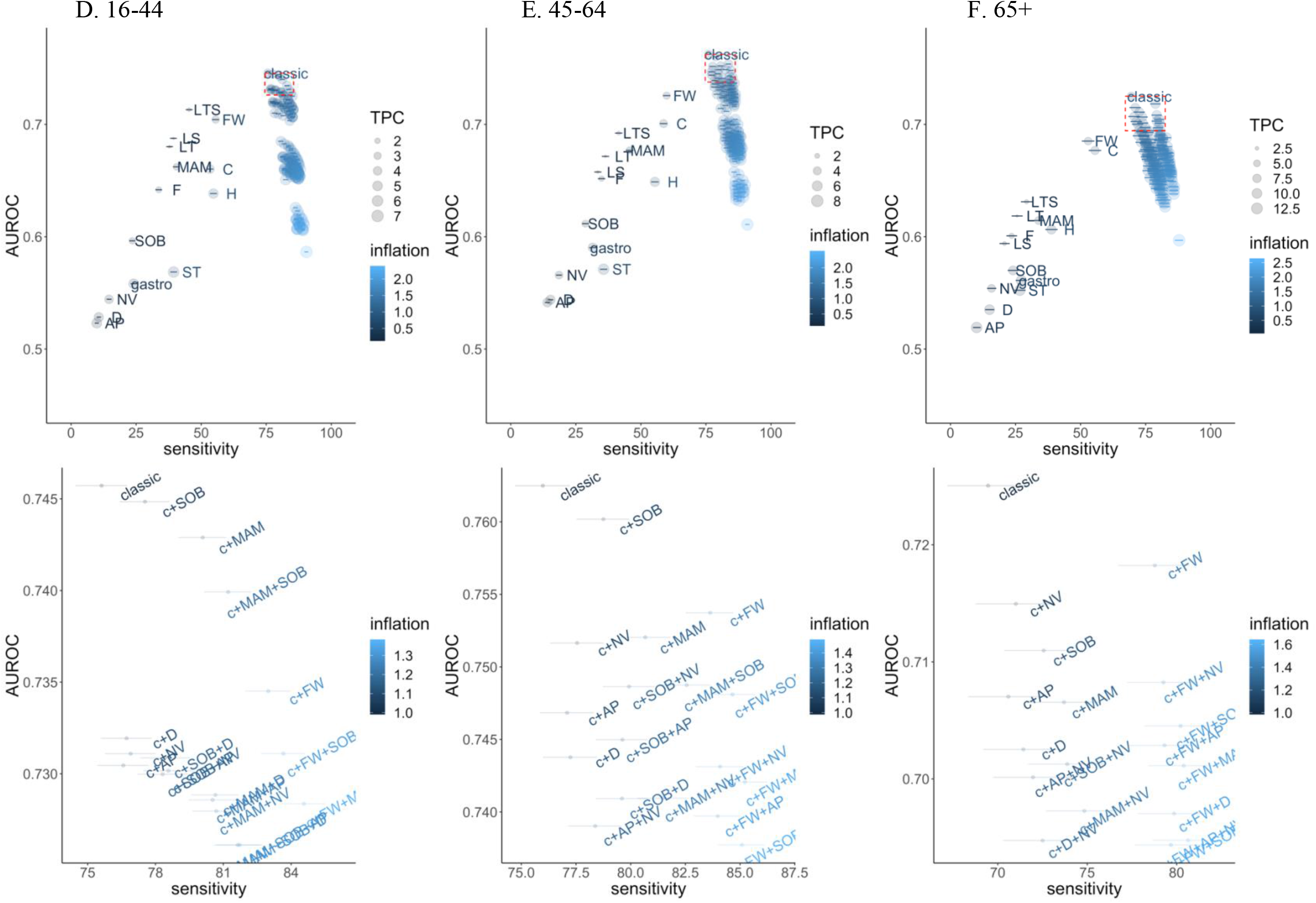
Performance of individual symptoms, as well as the classic four symptoms (cough, fever, loss of taste/smell), classic plus all possible combinations of 1/2/3/4 symptoms, and any of the 12 named symptoms, in predicting SARS-CoV-2 positivity in those with evidence of symptoms in terms of sensitivity and overall accuracy (AUROC). Note: Bottom row is an expanded version of the top right corner of the top row panels (red box, AUROC >90^th^ quantile, sensitivity > sensitivity of combination of classic 4 symptoms). Inflation (relative numbers reporting these symptoms compared to classic symptoms) and tests per positive case (TPC) are also included in the visualisation. TPC=1/positive predictive value. By definition, as the number of symptoms increases, sensitivity also increases. Note: abbreviations: c – classic, Fever - F, Headache - H, Muscle ache/myalgia - MAM, Weakness/tiredness - FW, Nausea/vomiting - NV, Abdominal pain - AP, Diarrhoea - D, Sore throat - ST, Cough - C, Shortness of breath - SOB, Loss of taste - LT, Loss of smell – LS, Loss of taste or smell – LTS

**Figure S8.**
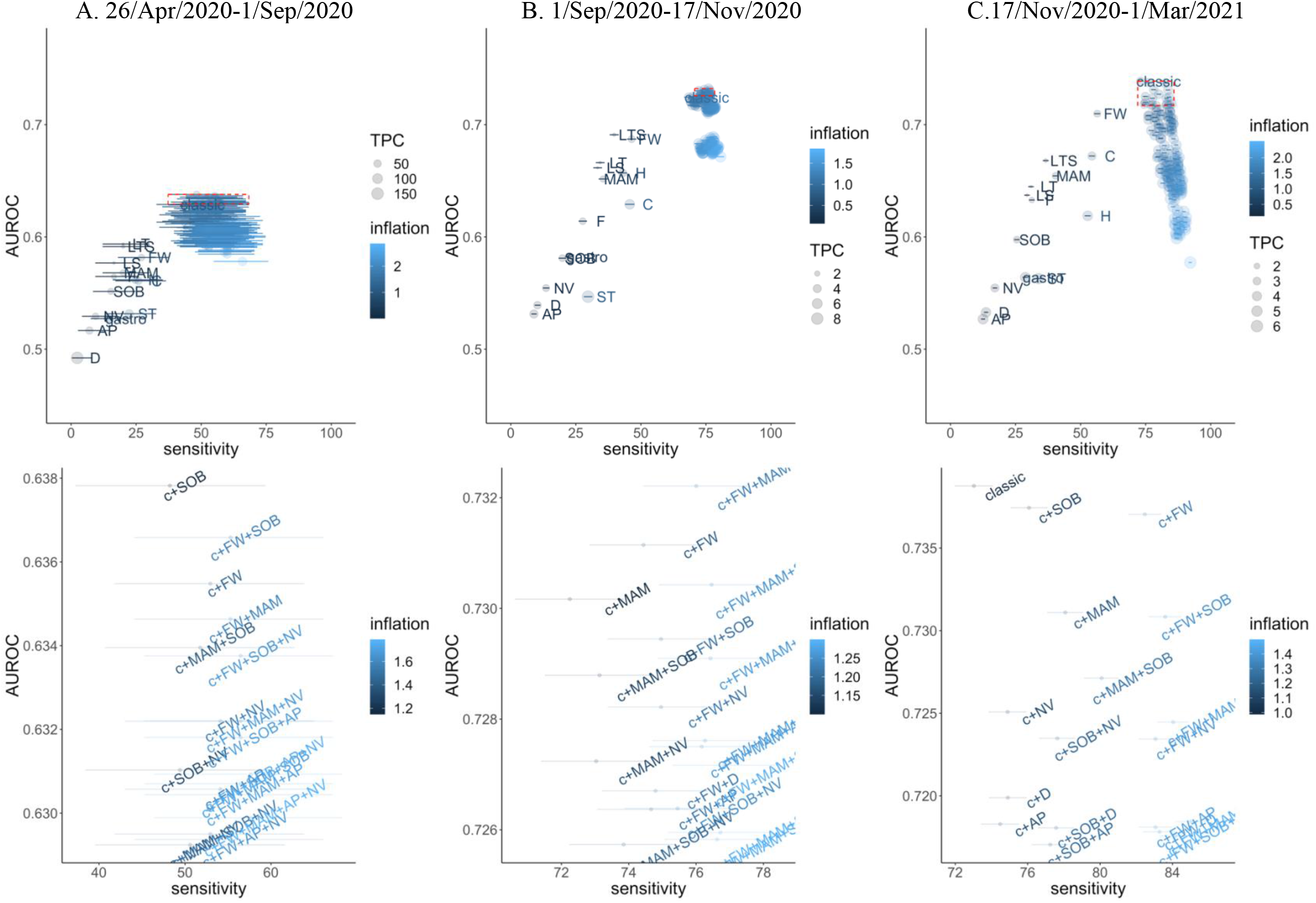
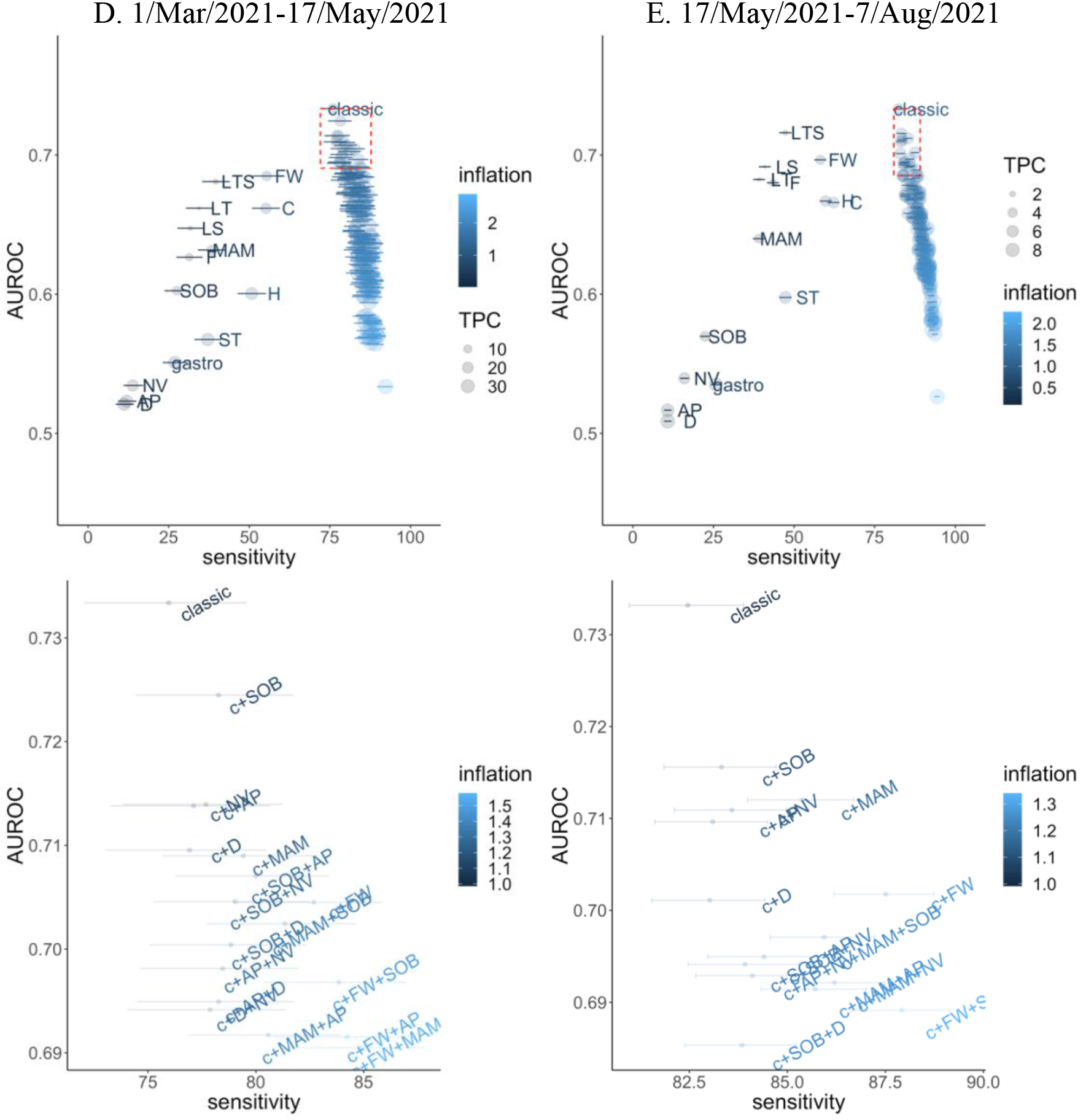
Performance of individual symptoms, as well as the classic four symptoms (cough, fever, loss of taste/smell), classic plus all possible combinations of 1/2/3/4 symptoms, and any of the 12 named symptoms, in predicting SARS-CoV-2 positivity in those with evidence of symptoms in terms of sensitivity and overall accuracy (AUROC). Note: Bottom row is an expanded version of the top right corner of the top row panels (red box, AUROC >90^th^ quantile, sensitivity > sensitivity of combination of classic 4 symptoms). Inflation (relative numbers reporting these symptoms compared to classic symptoms) and tests per positive case (TPC) are also included in the visualisation. TPC=1/positive predictive value. By definition, as the number of symptoms increases, sensitivity also increases. Note: abbreviations: c – classic, Fever - F, Headache - H, Muscle ache/myalgia - MAM, Weakness/tiredness - FW, Nausea/vomiting - NV, Abdominal pain - AP, Diarrhoea - D, Sore throat - ST, Cough - C, Shortness of breath - SOB, Loss of taste - LT, Loss of smell – LS, Loss of taste or smell – LTS

**Figure S9.**
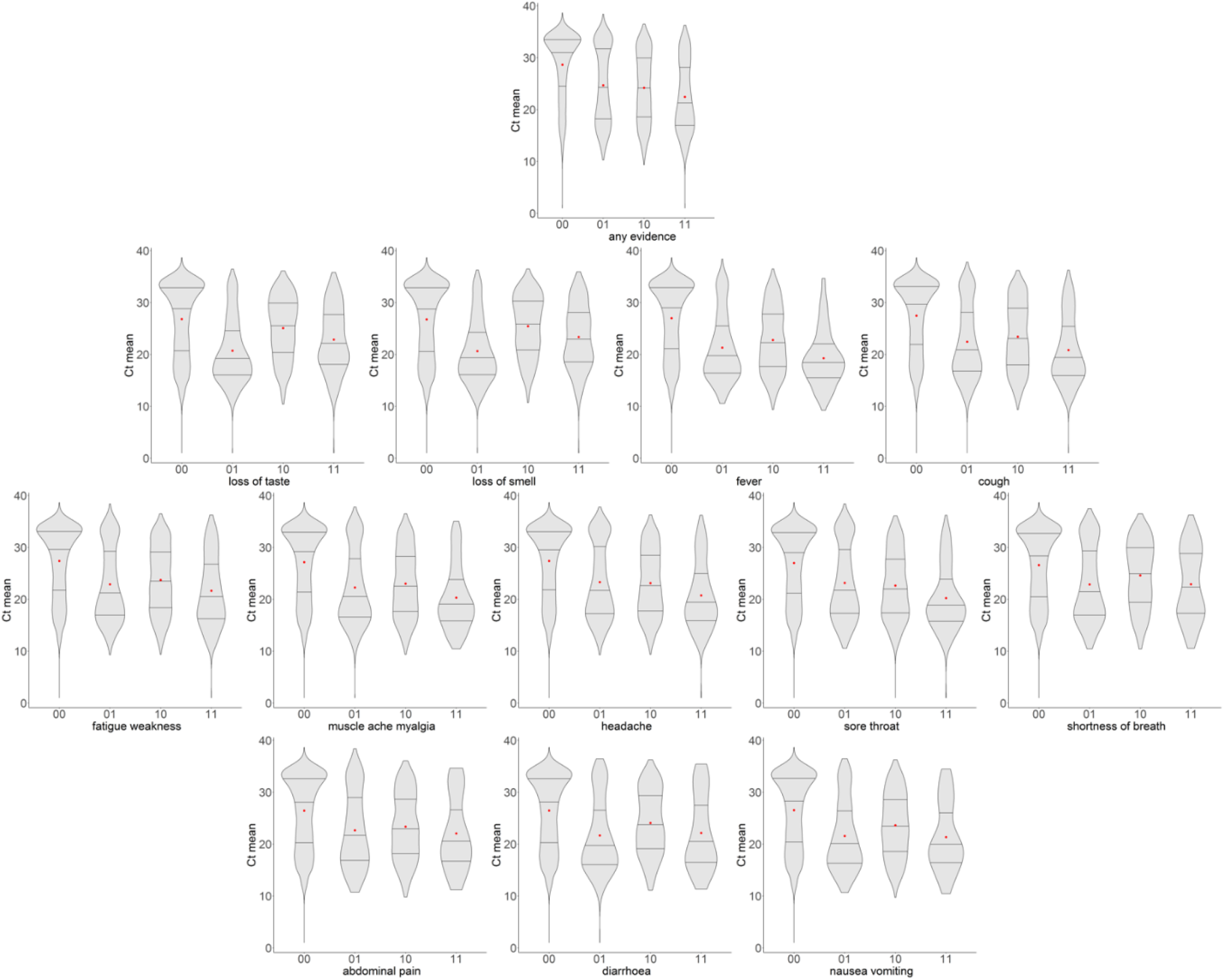
Distribution of Ct for each of the 12 symptoms in positive episodes with symptoms absent at all visits within 35 days of the index positive (00), absent at the index positive, but present at least one subsequent visit (01), present at the index positive, but absent at subsequent visits (10), present initially and at least one subsequent visit (11)

**Figure S10.**
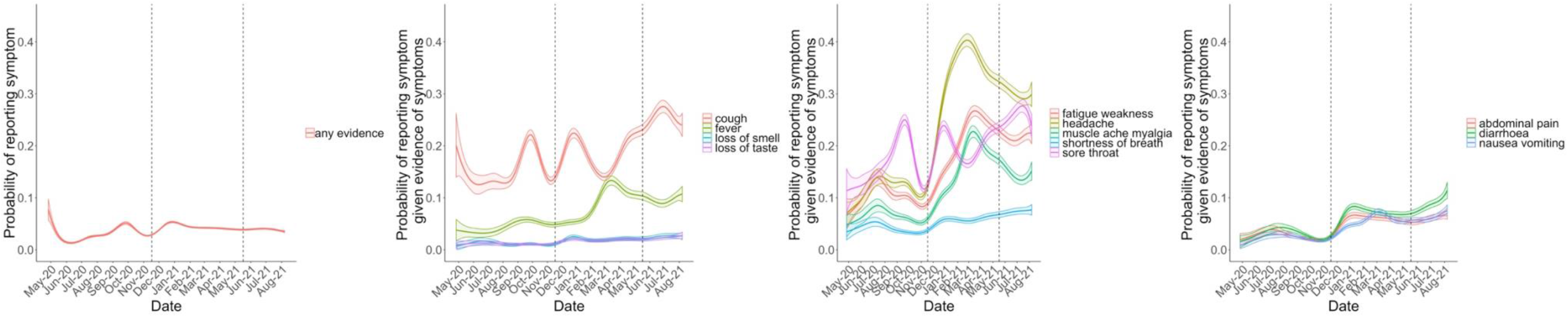
Probability of reporting specific symptoms at negative-visits with any evidence of symptoms before the final exclusion of negative visits where visit date is within [-7,+14] days of date of either first or second vaccination Note: adjusted for sex, age and ethnicity (reference category age 45, male, white ethnicity). Vaccination programme started 8 December 2020, but increased in magnitude in January 2021.

**Table S1.** Negative visits; summary of exclusions 0. Visits where PCR test negative
1. **<u>From -90 days before</u>** the first antibody positive test in the study prior to **vaccination**, where antibody results are likely to represent previous undetected infection;
2. **<u>From -35 days before</u>** the first positive onwards from individuals who ever tested PCR positive in the study or in the linked English testing programme (to avoid ongoing long COVID symptoms,^2^ and COVID-19-related symptoms shortly before the positive test);
3. **<u>From -35 days before</u>** any self-reported positive swab test result onwards (for the same reason; reflecting the fact that individuals may have obtained tests elsewhere)
4. From a small number of individuals who reported either loss of taste or loss of smell at their first study visit and had no national testing programme result within [-21,+21] days (all before 1 July 2020), given the high specificity of this symptom for COVID-19 infection, the fact that it would have been impossible for these individuals to get an external test and the potential for subsequent symptoms to represent long COVID
5. Where participants reported self-isolating OR contact with **<u>definite</u>** positives in the preceding 28 days (since these individuals have much higher risk of SARS-CoV-2 infection which may not have been detected) and the **<u>previous and the next visit</u>** (because of higher risk of unidentified positivity, and because they may have been contact traced through the national training programme they may be more likely to report symptoms through recall bias, regardless of status)
6. Occurring within [-7,+14 days] of either first or second vaccination date^3^, to avoid the inclusion of common symptoms caused by vaccination in the test-negative comparator group and to reflect the possibility of small inaccuracies in reported date of vaccination for some participants

| Negative visits excluding | With and without reported symptoms |  | With reported symptoms |  |
| --- | --- | --- | --- | --- |
|  | Visits (% from first row) | Participants (% from first row) | Visits (% from first row) | Participants (% from first row) |
| 0 | 5,095,824 | 489,804 | 209,281 | 142,513 |
| 1 | 4,981,365 (98%) | 482,963 (99%) | 202,429 (97%) | 138,393 (97%) |
| 1+2 | 4,795,502 (94%) | 471,641 (96%) | 189,041 (90%) | 130,242 (91%) |
| 1+2+3 | 4,708,088 (92%) | 464,232 (95%) | 183,380 (88%) | 126,777 (89%) |
| 1+2+3+4 | 4,702,276 (92%) | 463,638 (95%) | 182,128 (87%) | 126,183 (89%) |
| 1+2+3+4+5 | 4,172,632 (82%) | 458,510 (94%) | 149,822 (72%) | 108,773 (76%) |
| 1+2+3+4+5+6 | 3,806,692 (75%) | 457,215 (93%) | 130,612 (62%) | 97,186 (68%) |
| Summary | **130,612/3,806,692 (3.4%) of negative visits reported symptoms**<br>**97,816/457,215 (21%) of participants in the negative cohort reported symptoms** |  |  |  |

**Table S2.**
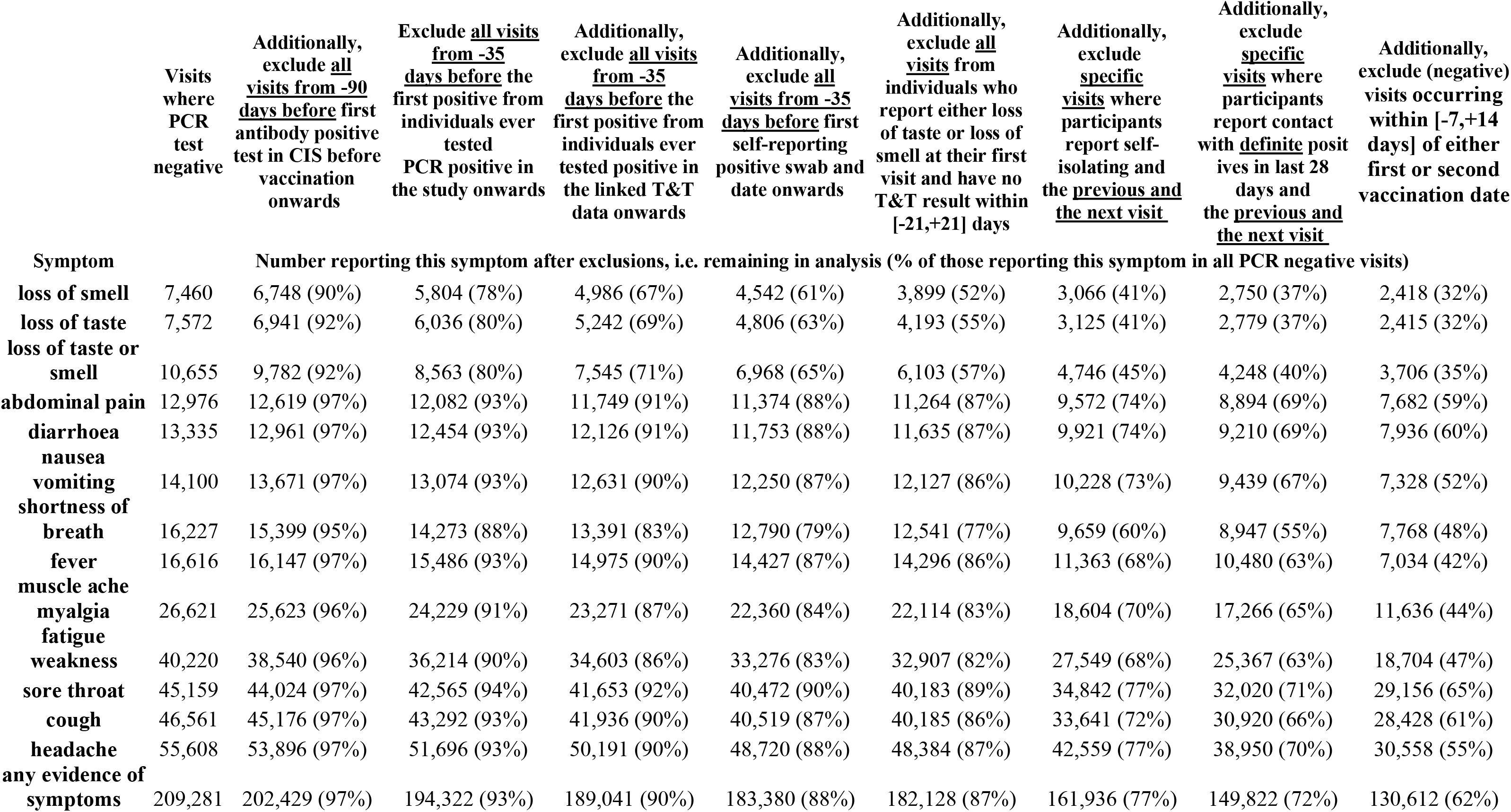
Numbers with each symptom in test-negative visits and percentage remaining from each as exclusion becomes stricter

**Table S3.**
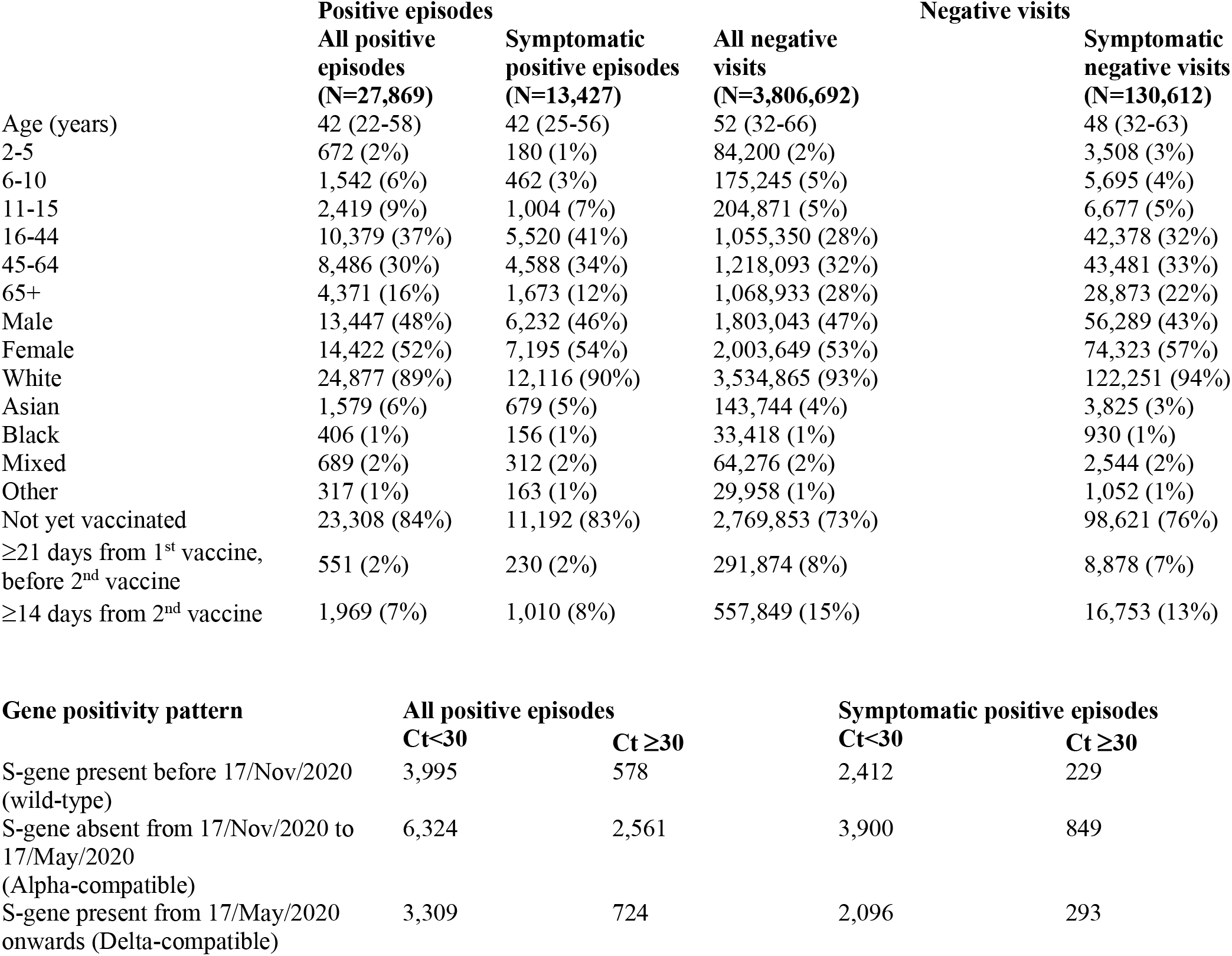
Characteristics of all and symptomatic positive episodes and all and symptomatic negative visits Note: showing median (IQR) or n (col %). Overall PPV of any evidence of symptoms predicting PCR positive episodes is 9%.

**Table S4.**
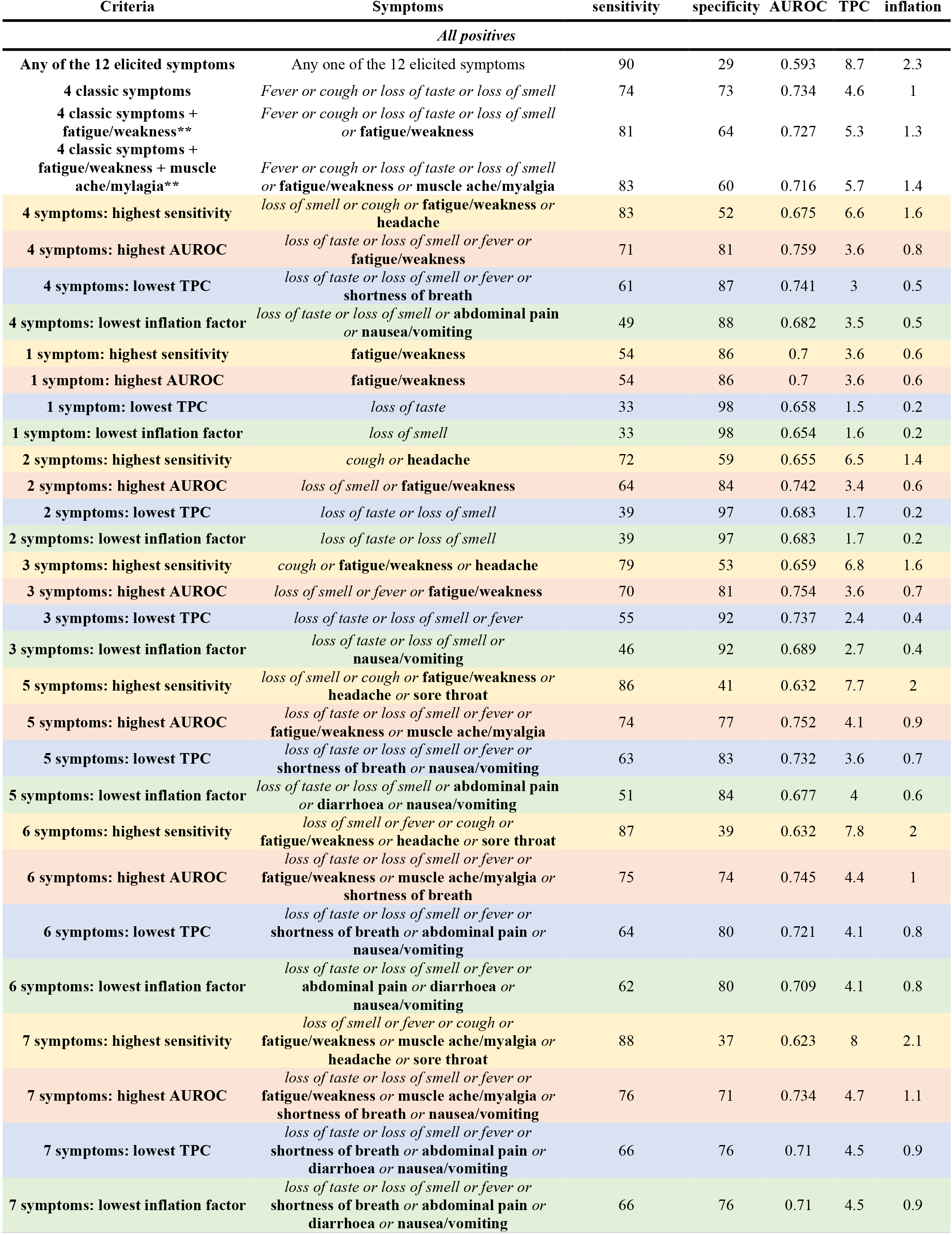
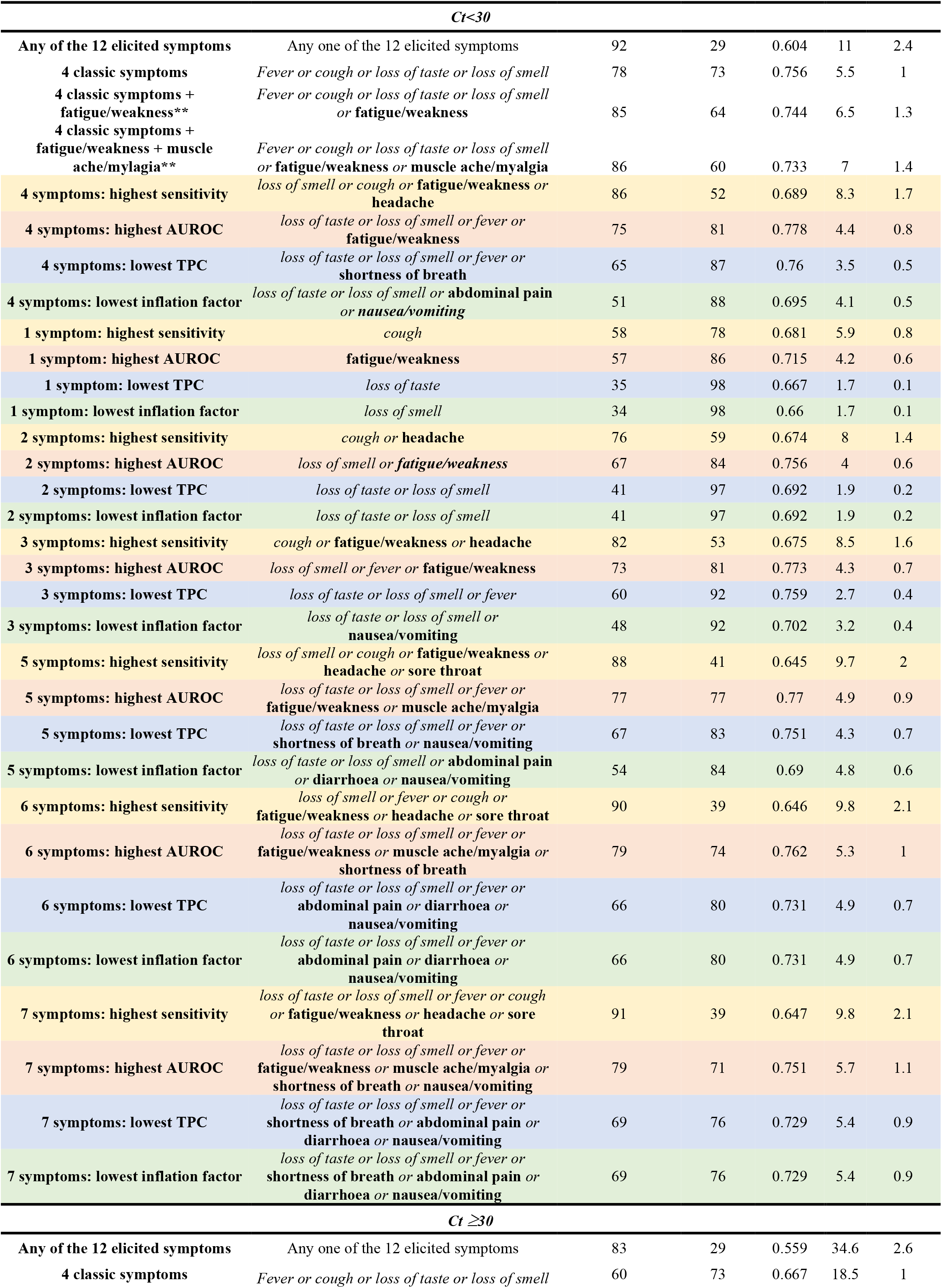
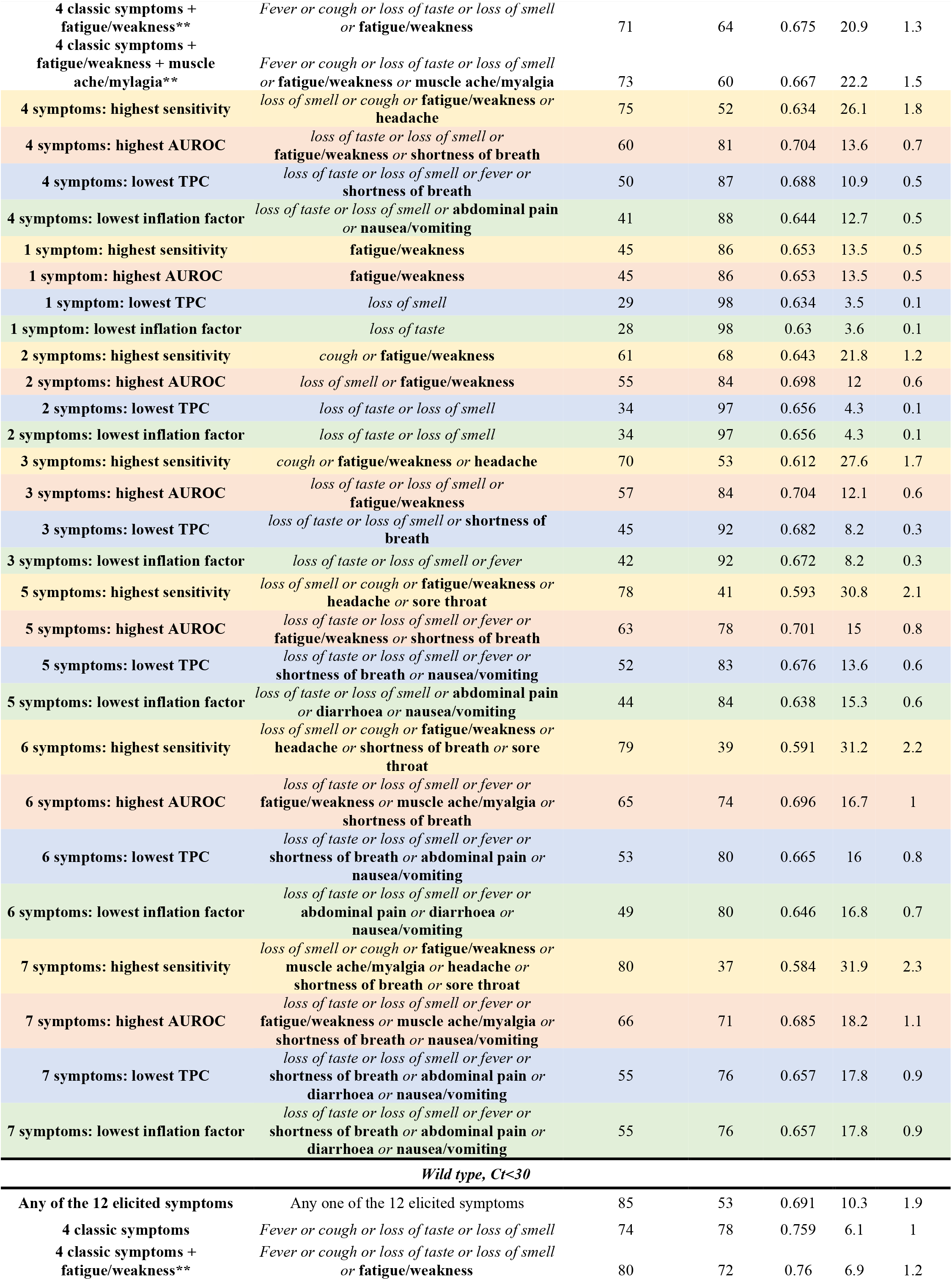
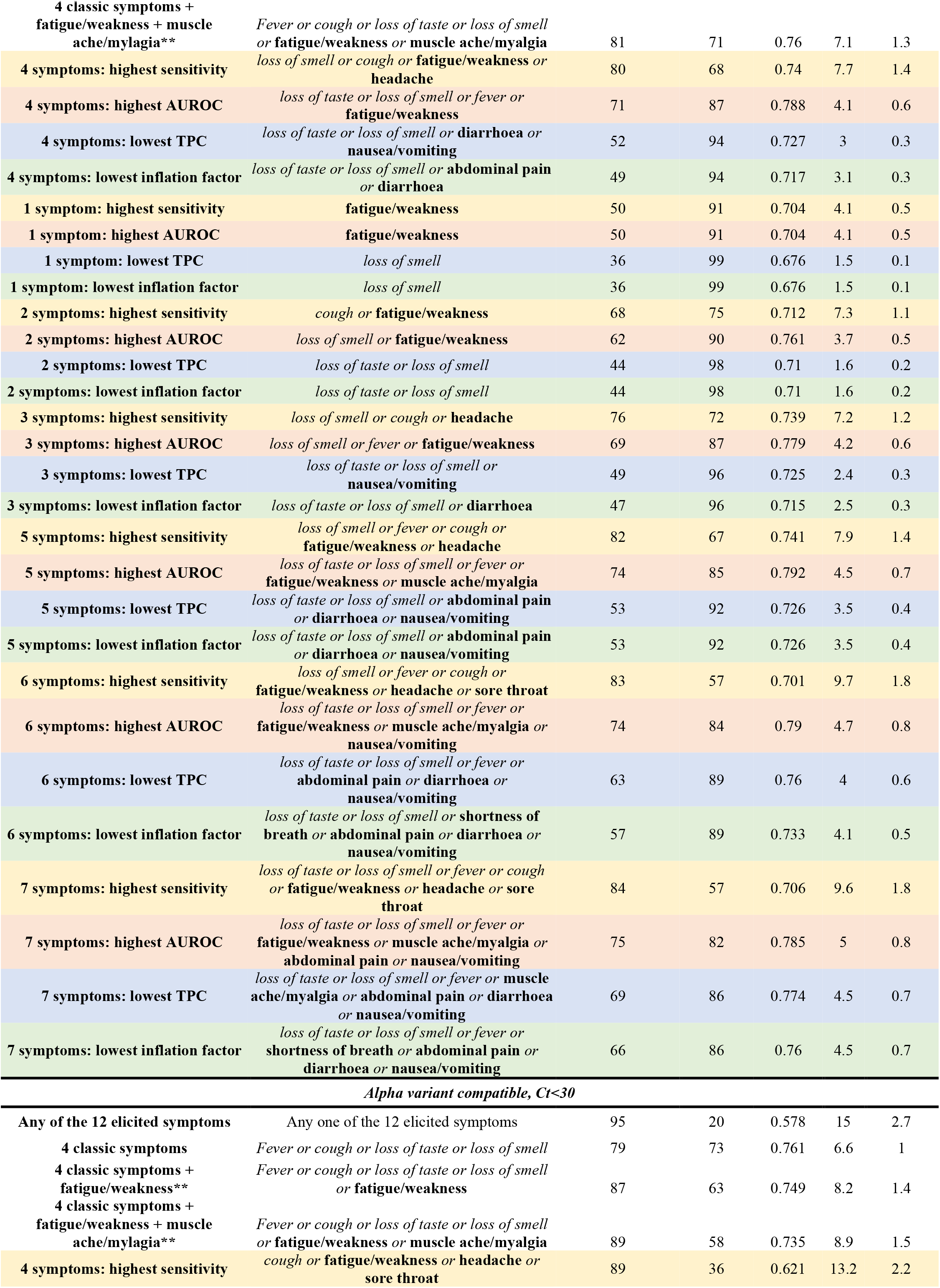
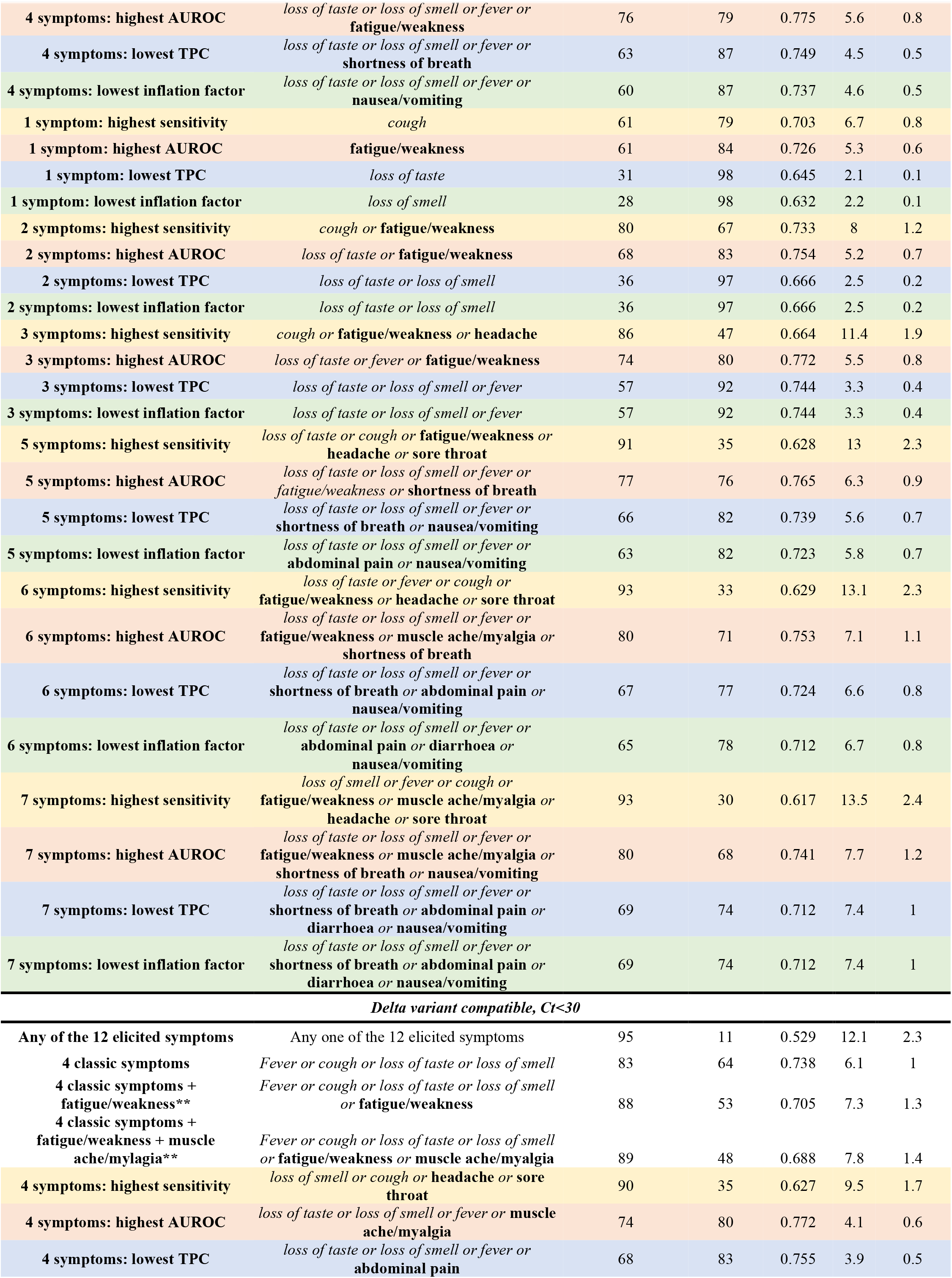
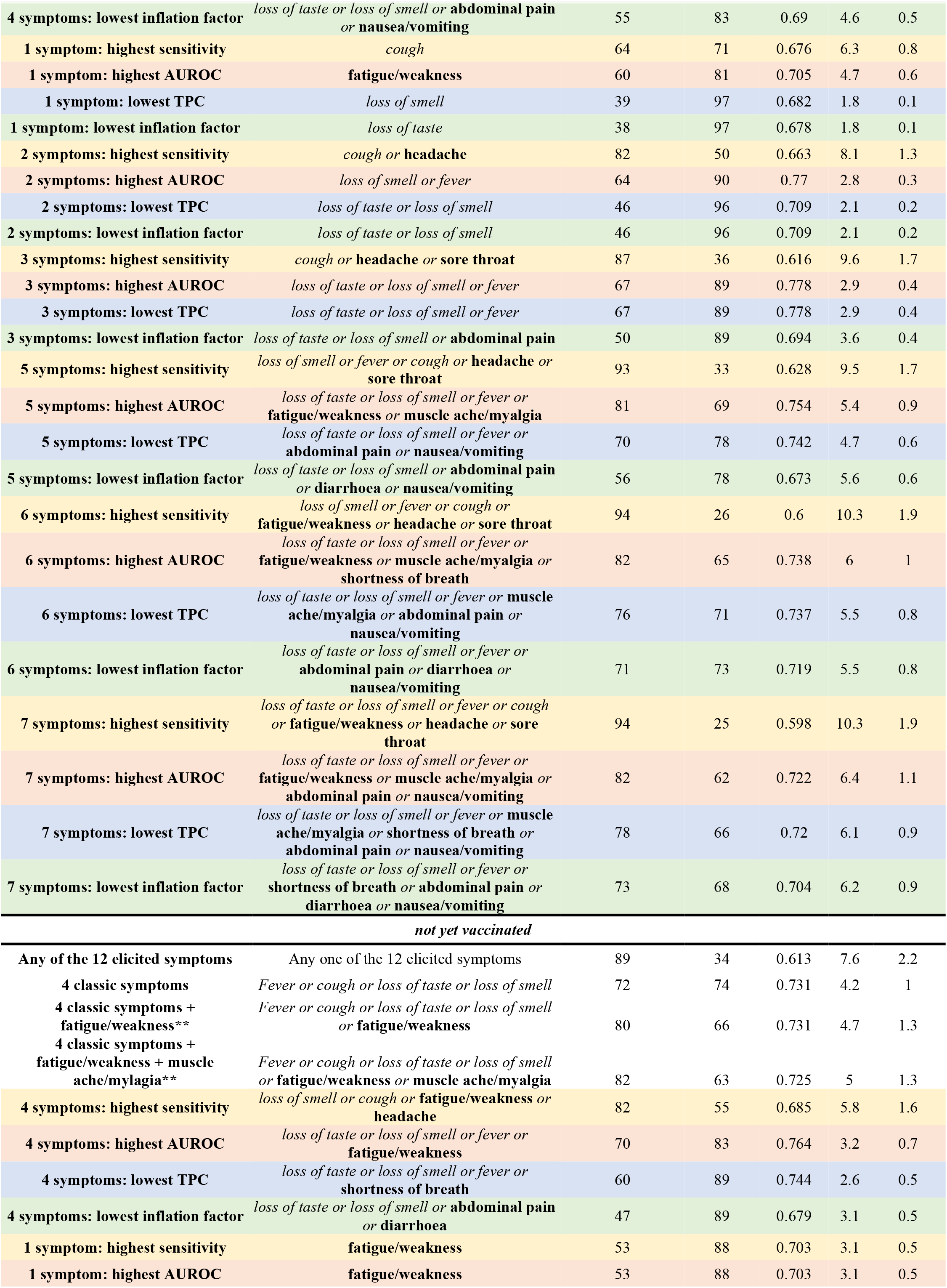
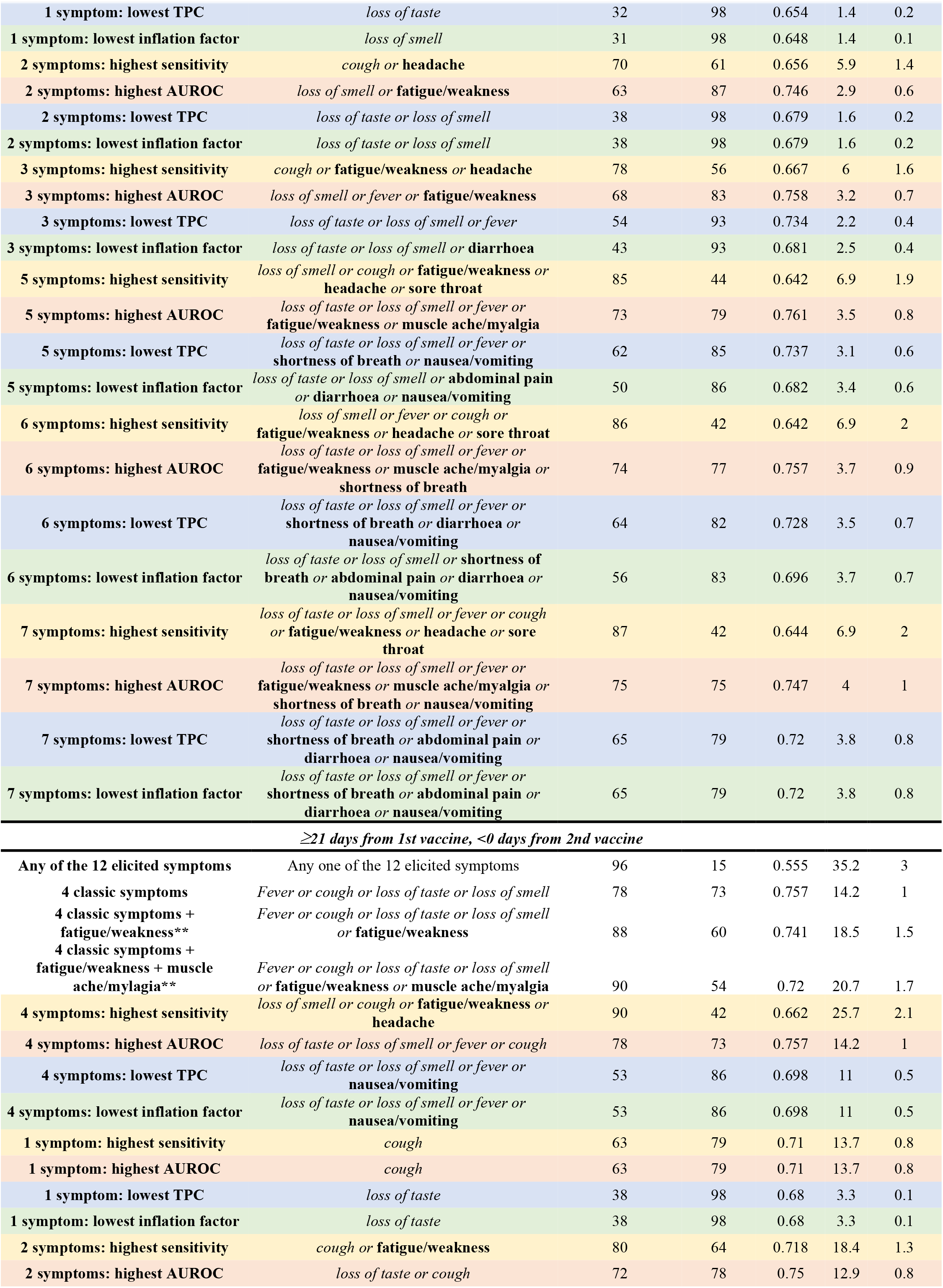
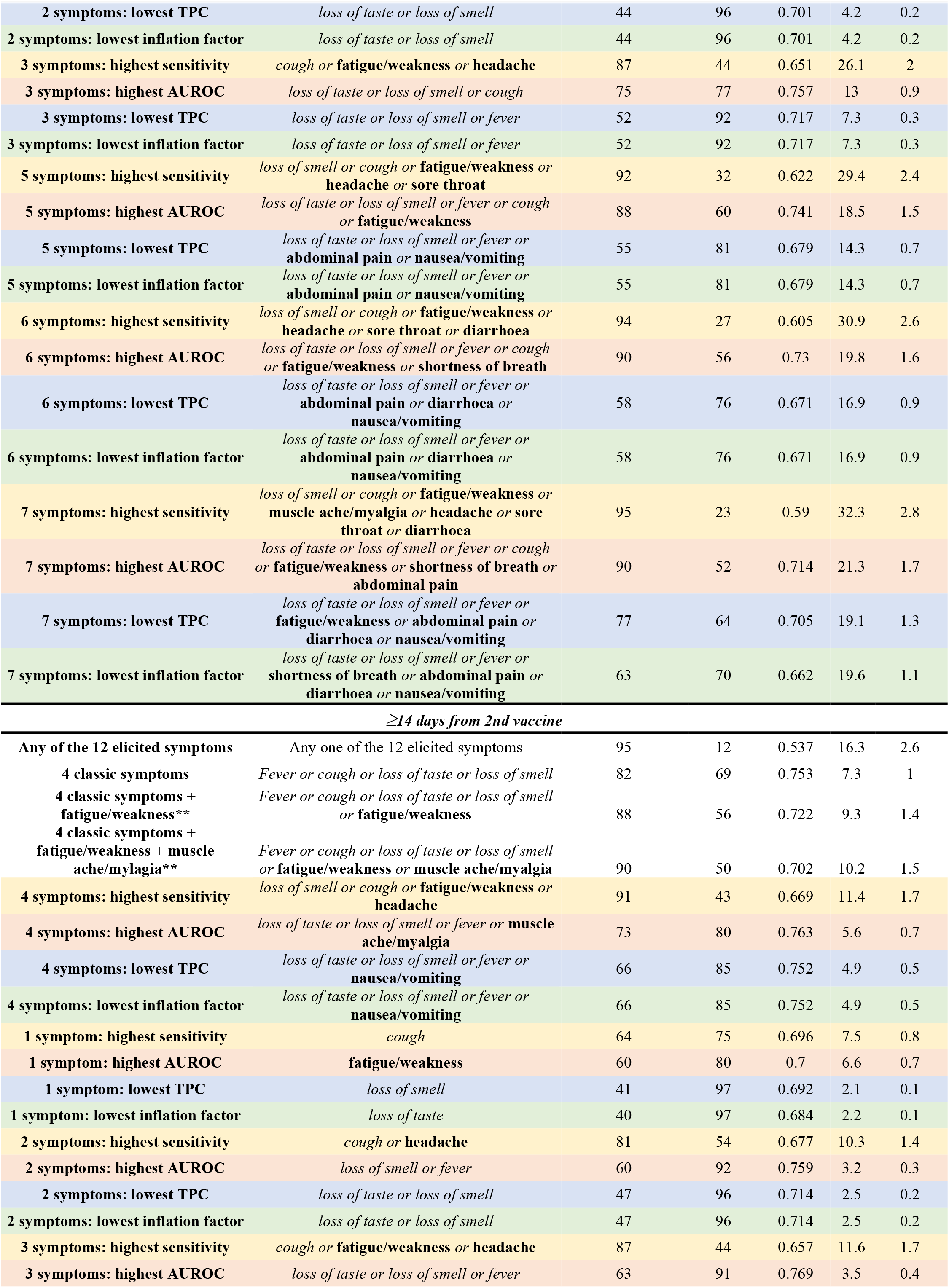
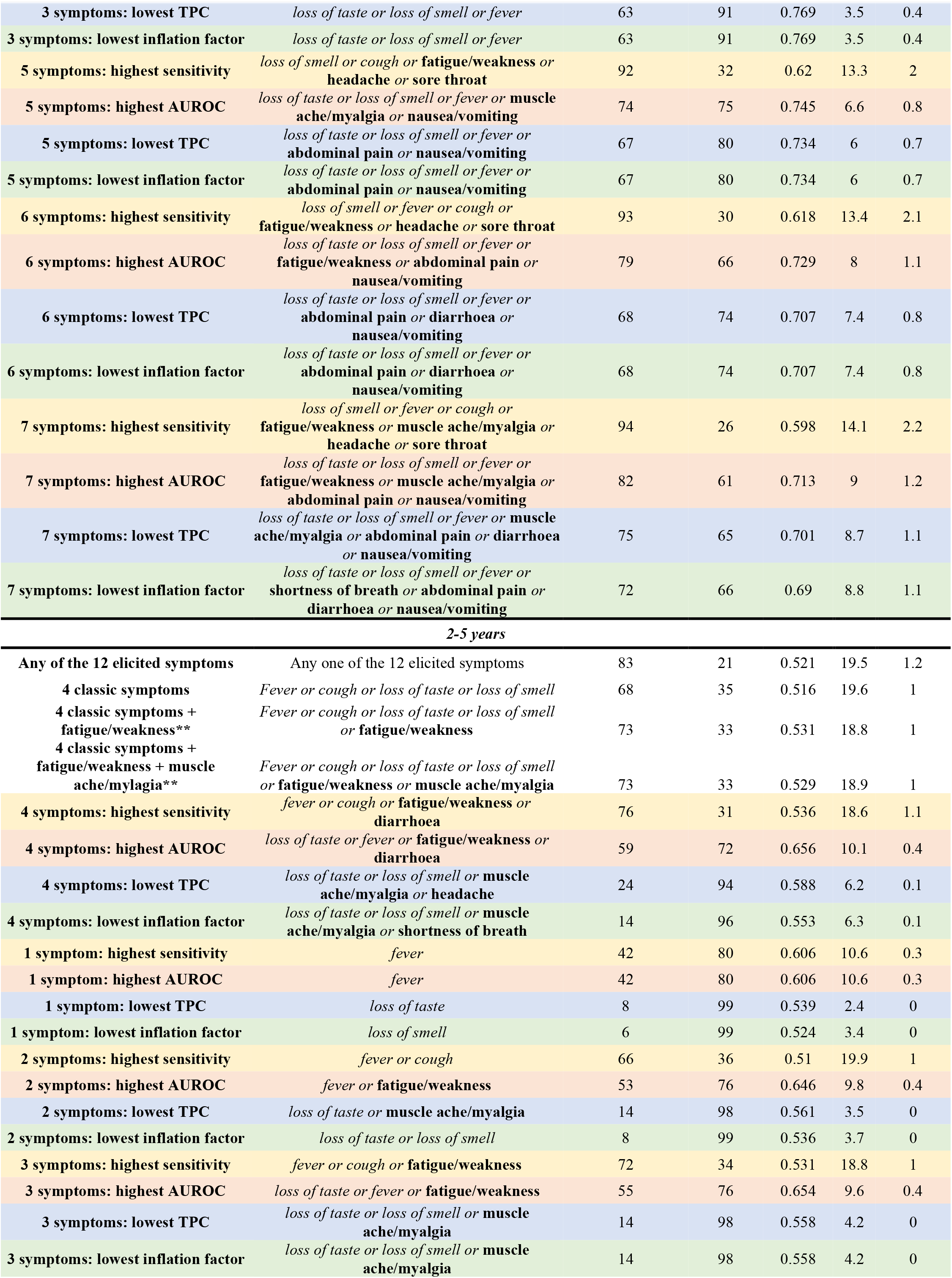
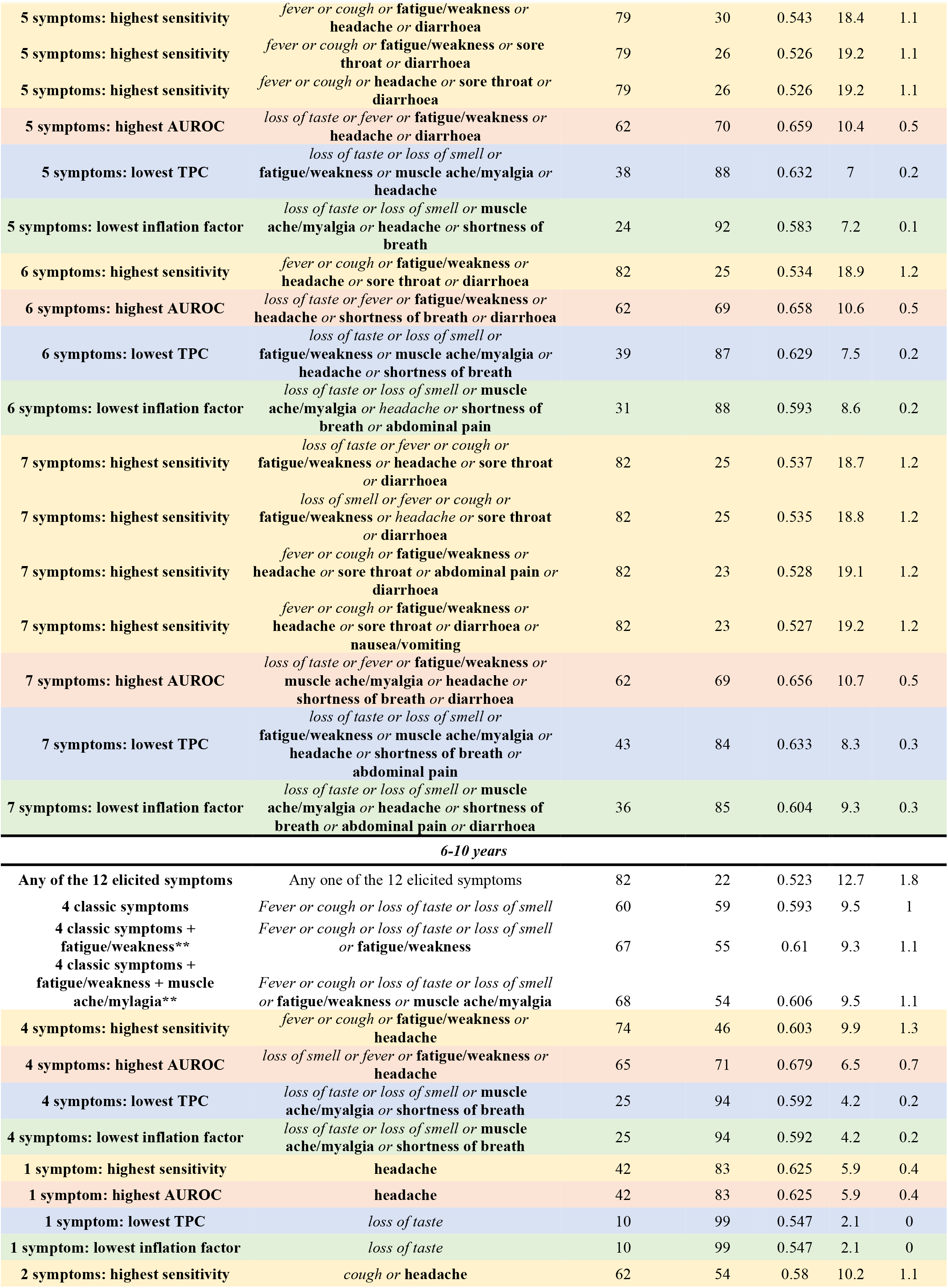
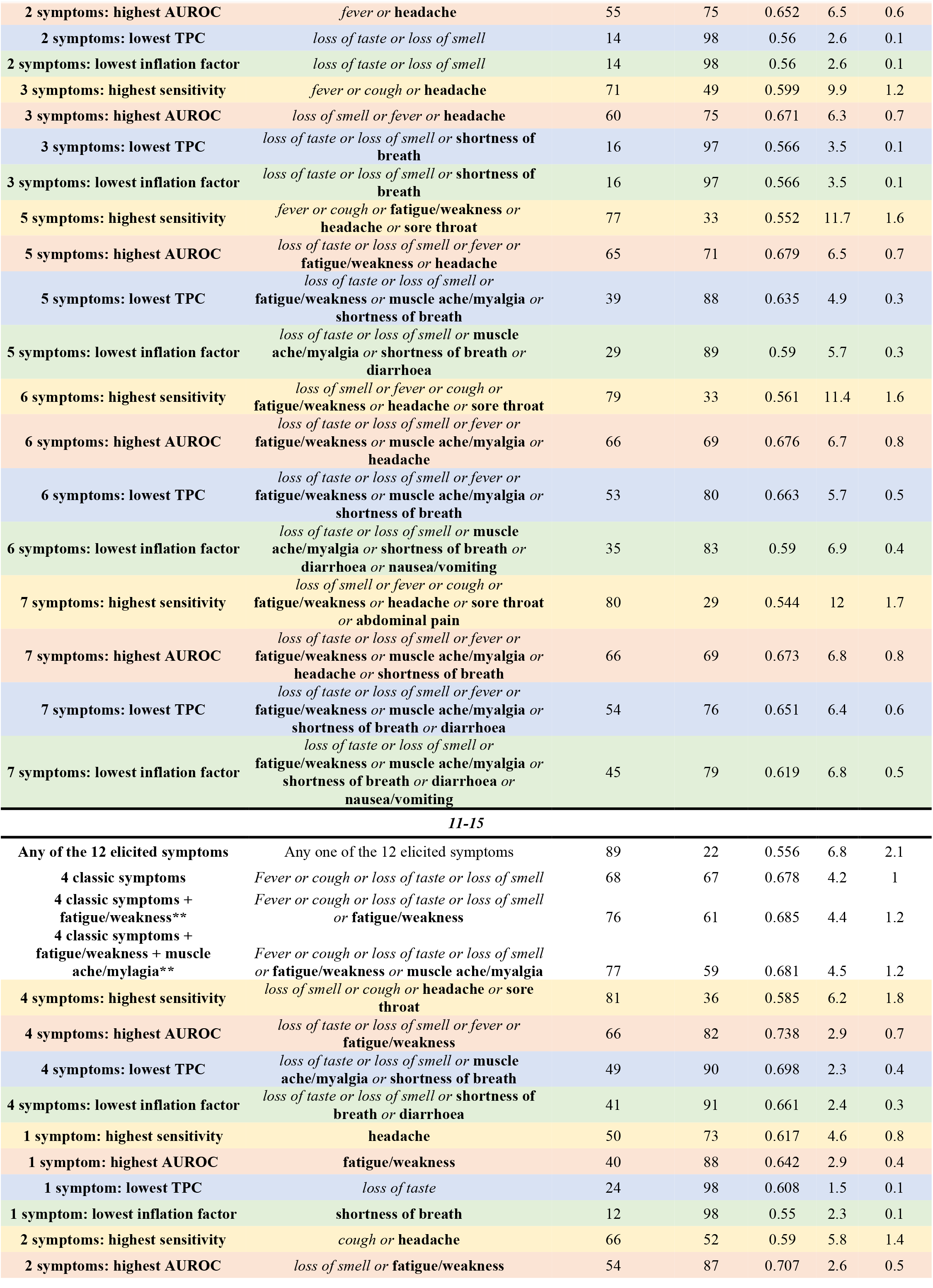
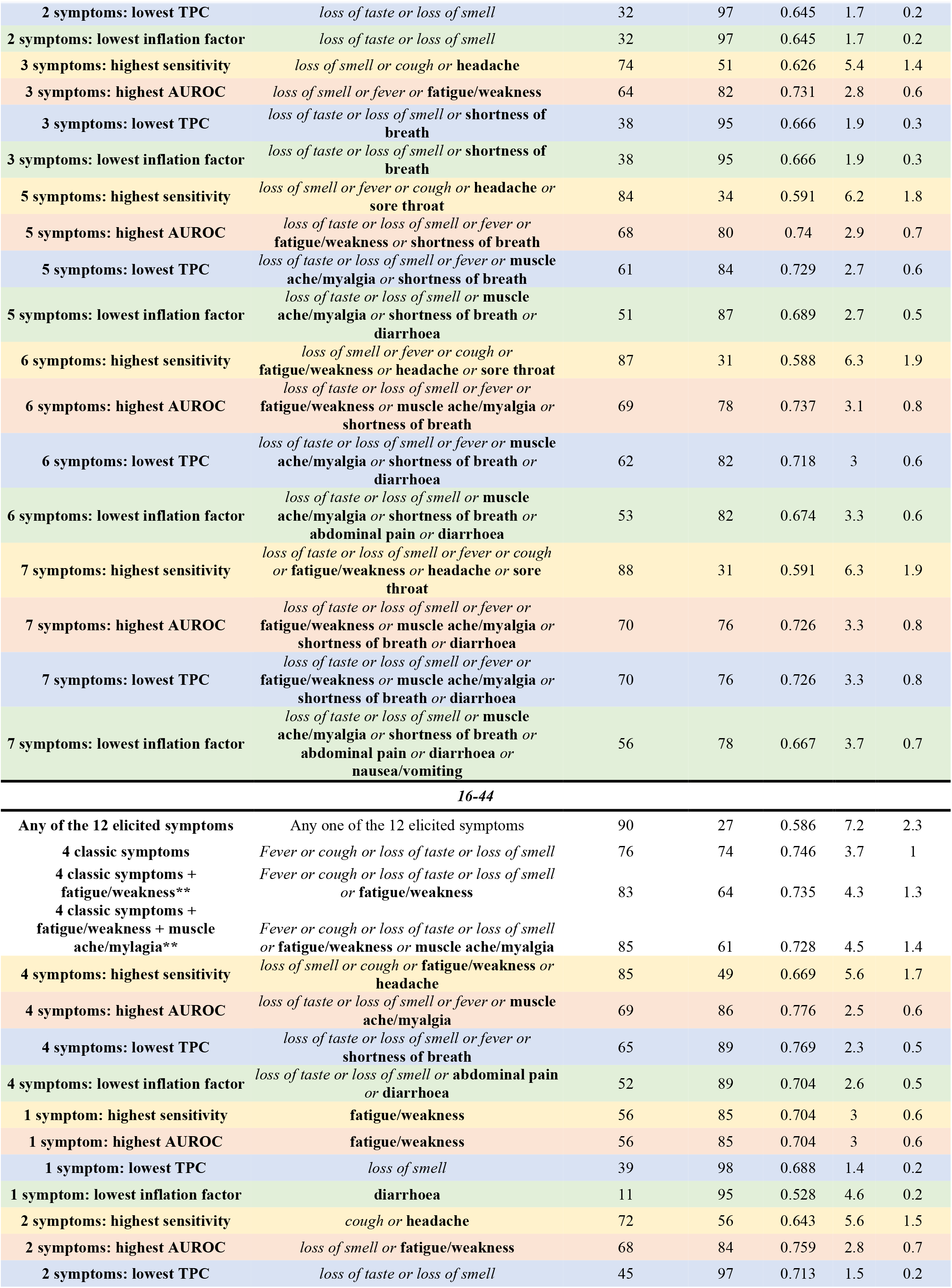
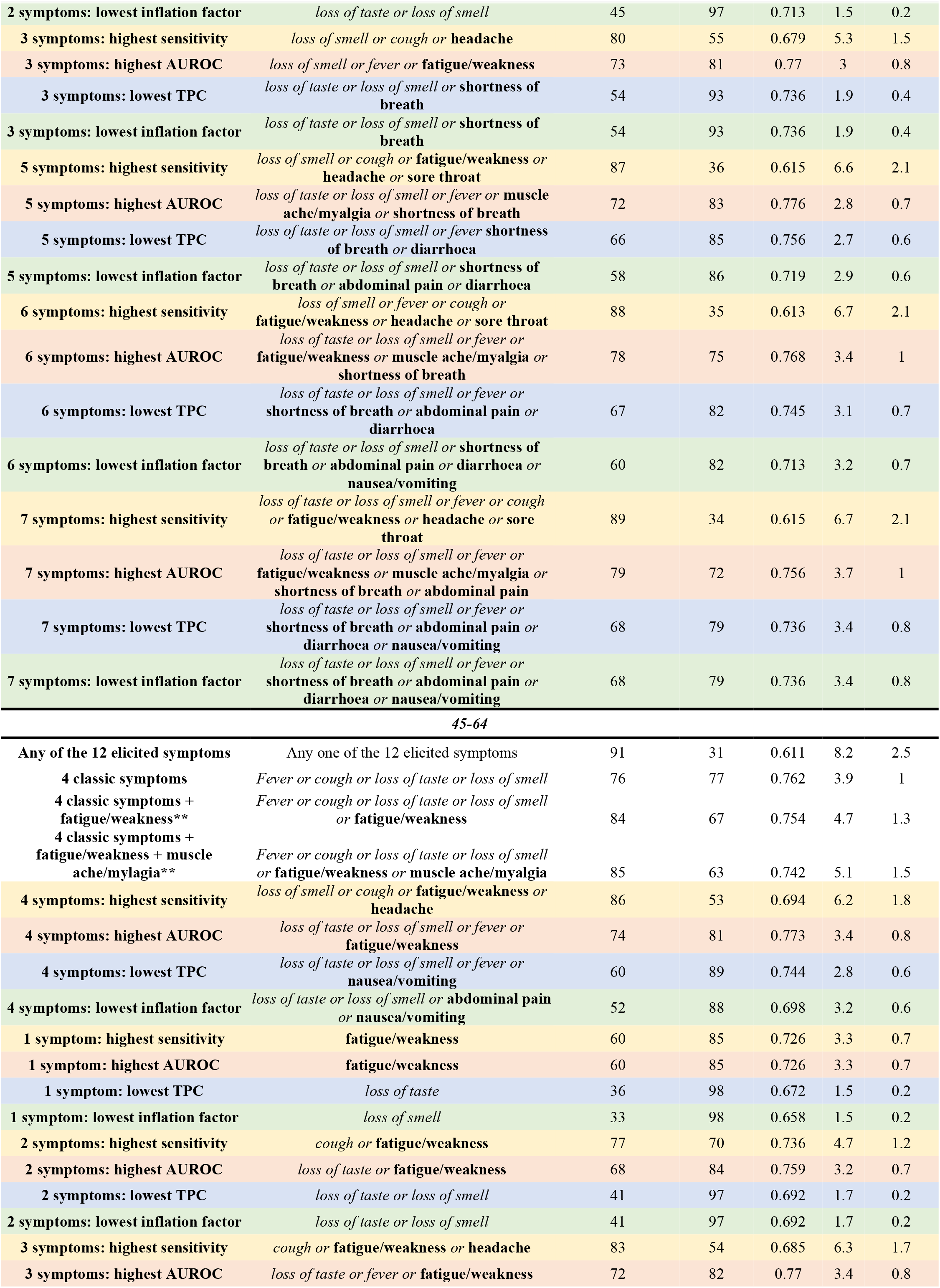
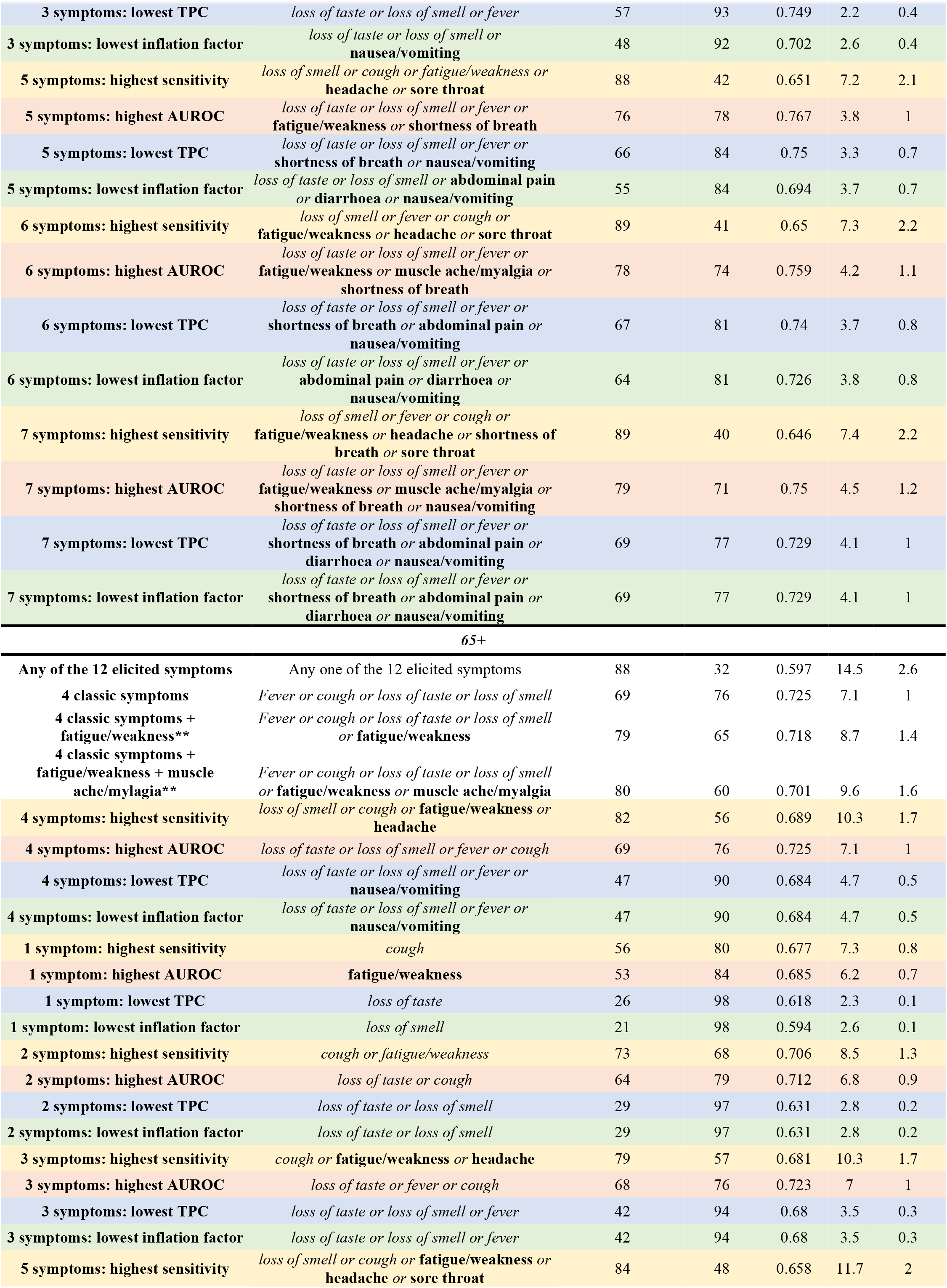
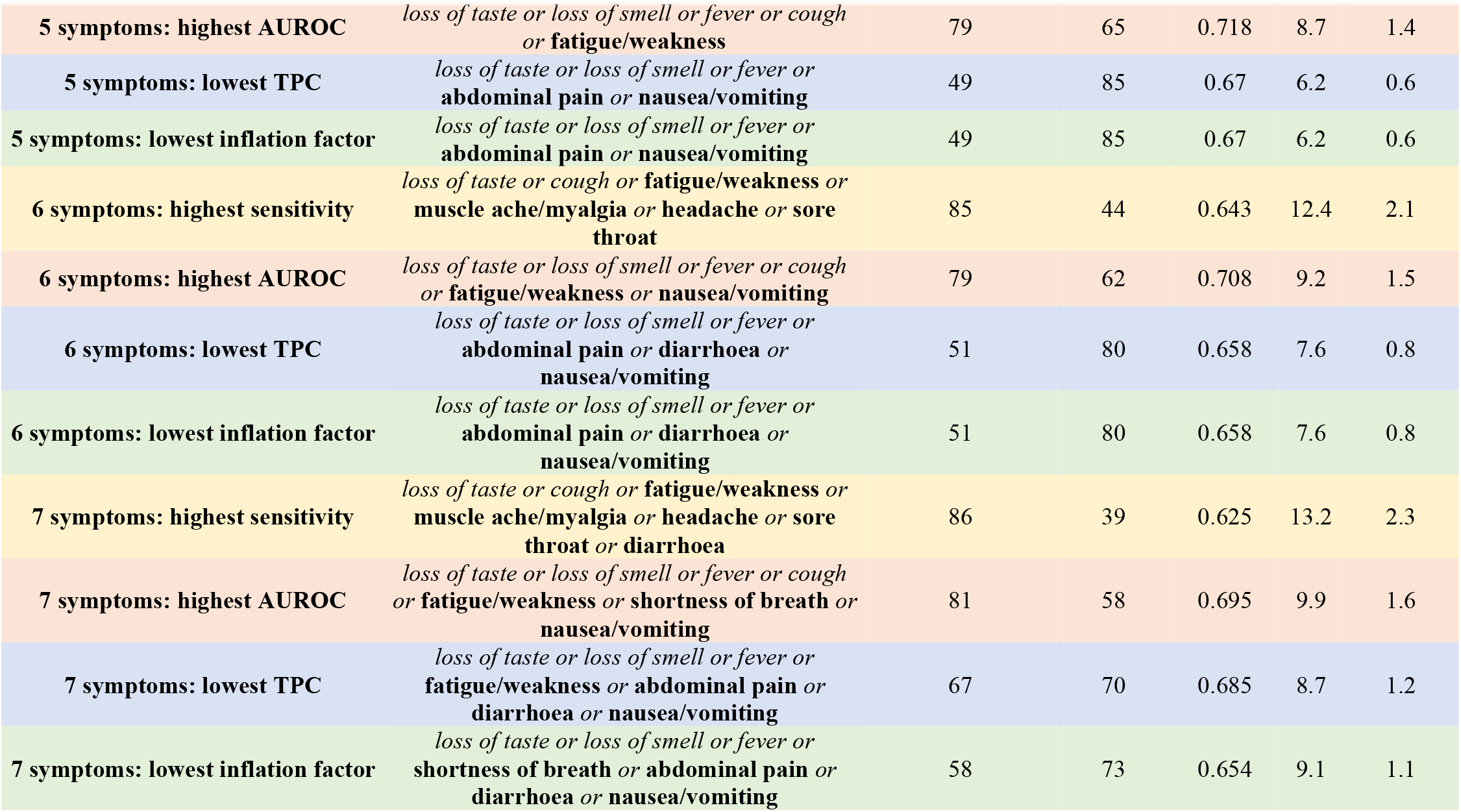
Sensitivity, specificity, test per case (TPC), AUROC and inflation factor for various symptom combinations across the whole study period and split by Ct valuet, variant, vaccination status and age Note: question about fatigue/weakness asked as “weakness/tiredness” to participants. Classic symptoms italicised, other symptoms in bold. *TPC=1/PPV **See **Table S5**

**Table S5.** Sensitivity, test per case (TPC), AUROC and inflation factor for various symptom combinations in different groups of positive episodes vs test-negative visits Note: For any number of symptoms, the combinations achieving the highest sensitivity and highest AUROC are shown in bold, and optimizing both in grey shading and bold, excluding the combination of any of the 12 named symptoms, given they achieve the highest sensitivity by definition

| Subgroup | classic (4) | classic or fatigue/weakness (5) | classic or muscle ache/myalgia (5) | classic or headache (5) | classic or diarrhoea (5) | any of the 12 named symptoms (12) |
| --- | --- | --- | --- | --- | --- | --- |
| All positives | **74%/4.6/0.734/1** | **81%/5.3/0.727/1.3** | 78%/5.1/0.726/1.2 | **82%/6.3/0.686/1.5** | 75%/5.1/0.717/1.1 | 90%/8.7/0.593/2.3 |
| Ct<30 | **78%/5.5/0.756/1** | **85%/6.5/0.744/1.3** | 82%/6.2/0.747/1.2 | **86%/7.8/0.704/1.6** | 79%/6.2/0.738/1.1 | 92%/11/0.604/2.4 |
| Ct≥30 | 60%/18.5/0.667/1 | **71%/20.9/0.675/1.3** | 66%/20.6/0.663/1.2 | 71%/25.7/0.63/1.6 | 62%/21/0.653/1.2 | 83%/34.6/0.559/2.6 |
| Wild type, Ct<30 | 74%/6.1/0.759/1 | **80%/6.9/0.76/1.2** | 78%/6.5/0.763/1.1 | 80%/7.3/0.75/1.3 | 75%/6.5/0.753/1.1 | 85%/10.3/0.691/1.9 |
| Alpha variant compatible, Ct<30 | **79%/6.6/0.761/1** | **87%/8.2/0.749/1.4** | 84%/7.7/0.75/1.2 | 87%/10.6/0.684/1.8 | 80%/7.7/0.741/1.2 | 95%/15/0.578/2.7 |
| Delta variant compatible, Ct<30 | **83%/6.1/0.738/1** | **88%/7.3/0.705/1.3** | 86%/6.9/0.717/1.2 | **90%/8.1/0.679/1.4** | 84%/7/0.705/1.2 | 95%/12.1/0.529/2.3 |
| Not yet vaccinated | 72%/4.2/0.731/1 | **80%/4.7/0.731/1.3** | 77%/4.5/0.729/1.2 | **81%/5.7/0.691/1.5** | 74%/4.6/0.719/1.1 | 89%/7.6/0.613/2.2 |
| ≥21 days from 1st vaccine, <0 days from 2nd vaccine | **78%/14.2/0.757/1** | **88%/18.5/0.741/1.5** | 83%/18.1/0.73/1.3 | 85%/24/0.672/1.8 | 81%/17.2/0.735/1.2 | 96%/35.2/0.555/3 |
| ≥14 days from 2nd vaccine | **82%/7.3/0.753/1** | **88%/9.3/0.722/1.4** | 86%/8.7/0.731/1.3 | 88%/10.4/0.692/1.5 | 83%/8.9/0.716/1.2 | 95%/16.3/0.537/2.6 |
| 2-5y | 68%/19.6/0.516/1 | **73%/18.8/0.531/1** | 68%/19.6/0.516/1 | 72%/18.9/0.529/1 | 72%/19.3/0.523/1 | 83%/19.5/0.521/1.2 |
| 6-10y | 60%/9.5/0.593/1 | 67%/9.3/0.61/1.1 | 62%/9.6/0.593/1 | **74%/9.7/0.611/1.3** | 62%/10/0.583/1.1 | 82%/12.7/0.523/1.8 |
| 11-15y | 68%/4.2/0.678/1 | 76%/4.4/0.685/1.2 | **72%/4.3/0.679/1.1** | **80%/5.3/0.639/1.5** | 69%/4.4/0.668/1.1 | 89%/6.8/0.556/2.1 |
| 16-44y | **76%/3.7/0.746/1** | **83%/4.3/0.735/1.3** | **80%/4/0.743/1.2** | **84%/5.3/0.681/1.6** | 77%/4/0.732/1.1 | 90%/7.2/0.586/2.3 |
| 45-64y | **76%/3.9/0.762/1** | **84%/4.7/0.754/1.3** | 81%/4.6/0.752/1.2 | 83%/5.9/0.702/1.6 | 77%/4.5/0.744/1.2 | 91%/8.2/0.611/2.5 |
| 65y+ | **69%/7.1/0.725/1** | **79%/8.7/0.718/1.4** | 74%/8.6/0.707/1.3 | 76%/9.5/0.694/1.5 | 71%/8.5/0.703/1.2 | 88%/14.5/0.597/2.6 |
| 26Apr2020-30Aug2020 | 44%/37.9/0.629/1 | **53%/45.2/0.635/1.5** | 48%/43.3/0.628/1.3 | 49%/51.7/0.609/1.5 | 44%/44.4/0.613/1.2 | 66%/70/0.578/2.8 |
| 1Sep2020-16Nov2020 | 68%/4.7/0.724/1 | **74%/5.1/0.731/1.2** | 72%/4.9/0.73/1.1 | **75%/5.4/0.719/1.3** | 69%/4.9/0.72/1.1 | 81%/7.2/0.671/1.8 |
| 17Nov2020-28Feb2021 | **73%/3.2/0.739/1** | **82%/3.7/0.737/1.3** | 78%/3.6/0.731/1.2 | **83%/4.7/0.671/1.7** | 75%/3.6/0.72/1.2 | 92%/6.2/0.577/2.5 |
| 1Mar2021-16May2021 | **76%/16.5/0.733/1** | **83%/21.4/0.705/1.4** | 79%/20.1/0.709/1.3 | 82%/27.1/0.646/1.8 | 77%/19.4/0.71/1.2 | 92%/38.4/0.534/2.8 |
| 17May2021-17Jul2021 | **82%/5/0.733/1** | **88%/6/0.702/1.3** | 85%/5.6/0.712/1.2 | **89%/6.6/0.673/1.4** | 83%/5.8/0.701/1.2 | 94%/9.7/0.526/2.2 |

| Subgroup | classic or shortness of breath (5) | classic or sore throat (5) | classic or abdominal pain (5) | classic or nausea/vomiting (5) |
| --- | --- | --- | --- | --- |
| **All positives** | 76%/4.9/0.729/1.1 | 79%/6.1/0.687/1.4 | 75%/5.1/0.718/1.1 | 75%/5/0.722/1.1 |
| **Ct<30** | 80%/5.9/0.749/1.1 | 83%/7.5/0.708/1.4 | 79%/6.1/0.74/1.1 | 80%/6/0.744/1.1 |
| **Ct≥30** | 64%/19.3/0.672/1.1 | 66%/25.5/0.624/1.5 | 62%/21/0.651/1.2 | 62%/20.6/0.655/1.1 |
| **Wild type, Ct<30** | 75%/6.4/0.757/1.1 | 78%/8.5/0.715/1.5 | 75%/6.5/0.754/1.1 | 76%/6.4/0.757/1.1 |
| **Alpha variant compatible, Ct<30** | 81%/7.2/0.754/1.1 | 84%/9.3/0.712/1.5 | 80%/7.7/0.74/1.2 | 81%/7.5/0.747/1.2 |
| **Delta variant compatible, Ct<30** | 84%/6.6/0.719/1.1 | 89%/7.9/0.683/1.4 | 84%/6.8/0.714/1.1 | 85%/6.8/0.716/1.1 |
| **Not yet vaccinated** | 75%/4.4/0.731/1.1 | 77%/5.6/0.684/1.4 | 74%/4.6/0.717/1.1 | 74%/4.5/0.722/1.1 |
| **≥21 days from 1st vaccine, <0 days from 2nd vaccine** | 80%/16.5/0.741/1.2 | 84%/19.5/0.718/1.5 | 79%/16.9/0.733/1.2 | 79%/16.5/0.735/1.2 |
| **≥14 days from 2nd vaccine** | 83%/8.4/0.729/1.2 | 87%/9.7/0.707/1.4 | 83%/8.4/0.73/1.2 | 83%/8.3/0.732/1.1 |
| **2-5y** | 68%/19.6/0.515/1 | 71%/20.2/0.505/1.1 | 68%/20.2/0.505/1 | 70%/19.9/0.51/1.1 |
| **6-10y** | 60%/9.6/0.59/1 | 65%/11.8/0.54/1.4 | 63%/10.4/0.575/1.1 | 63%/10.2/0.58/1.1 |
| **11-15y** | 70%/4.2/0.681/1 | 76%/5.4/0.63/1.4 | 70%/4.6/0.658/1.1 | 71%/4.6/0.663/1.1 |
| **16-44y** | 78%/3.8/0.745/1.1 | 81%/5.2/0.685/1.5 | 77%/4.1/0.73/1.1 | 77%/4.1/0.731/1.1 |
| **45-64y** | 79%/4.2/0.76/1.1 | 81%/5.4/0.714/1.5 | 77%/4.4/0.747/1.1 | 78%/4.3/0.752/1.1 |
| **65y+** | 73%/8.2/0.711/1.2 | 74%/9.2/0.693/1.4 | 71%/8.1/0.707/1.2 | 71%/7.8/0.715/1.1 |
| **26Apr2020-30Aug2020** | 48%/39.8/0.638/1.2 | 53%/52.2/0.615/1.7 | 45%/43.1/0.62/1.2 | 46%/41.4/0.627/1.2 |
| **1Sep2020-16Nov2020** | 70%/4.8/0.725/1.1 | 72%/6.3/0.683/1.4 | 69%/4.9/0.72/1.1 | 70%/4.9/0.723/1.1 |
| **17Nov2020-28Feb2021** | 76%/3.4/0.737/1.1 | 78%/4.2/0.69/1.4 | 74%/3.6/0.718/1.2 | 75%/3.5/0.725/1.1 |
| **1Mar2021-16May2021** | 78%/18.2/0.724/1.1 | 81%/23.1/0.683/1.5 | 78%/19.1/0.714/1.2 | 77%/19/0.714/1.2 |
| **17May2021-17Jul2021** | 83%/5.5/0.716/1.1 | 88%/6.5/0.677/1.4 | 83%/5.6/0.71/1.1 | 84%/5.6/0.711/1.1 |

**Table S6.**
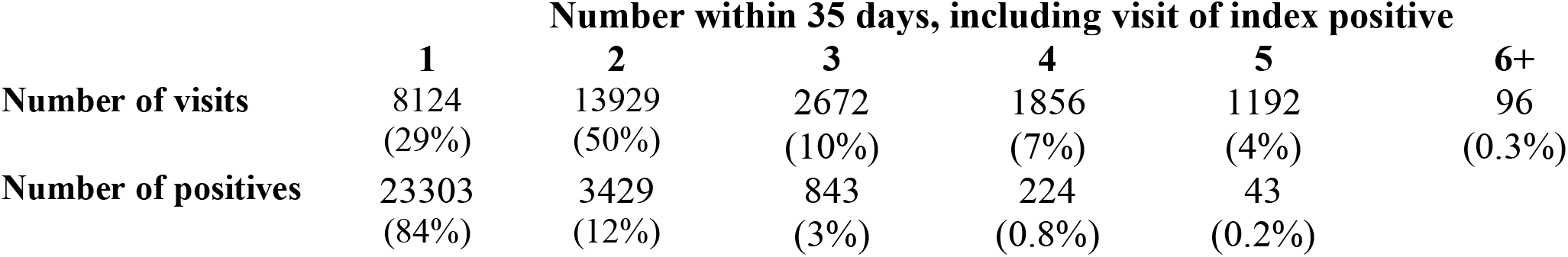
Number of visits and number of positive tests contributing to the analysis of symptoms reported in positive episodes (N=27,869)

|  | **Number within 35 days, including visit of index positive** |  |  |  |  |  |
| --- | --- | --- | --- | --- | --- | --- |
|  | **1** | **2** | **3** | **4** | **5** | **6+** |
| **Number of visits** | 8124<br>(29%) | 13929<br>(50%) | 2672<br>(10%) | 1856<br>(7%) | 1192<br>(4%) | 96<br>(0.3%) |
| **Number of positives** | 23303<br>(84%) | 3429<br>(12%) | 843<br>(3%) | 224<br>(0.8%) | 43<br>(0.2%) |  |

**Table S7.** Summary of number of symptoms among positive episodes present at index positive and ever present at any future visits within [0,35] days of the index positive, including at visits with negative or missing results (both absent, both present, absent then present, present then absent for each symptom). Note: symptoms are ordered from highest to lowest percentage of positive episodes where the symptom was absent at index positive, but present at at least one subsequent visit in the next 35 days

| Symptom<br>n(%) | Absent at both<br>index positive and all future visits<br>within 35 days | Absent at<br>index positive,<br>present at at least one<br>subsequent visit | Present at index positive,<br>absent at all future visits<br>within 35 days | Present at both index positive and at least one<br>subsequent visit |
| --- | --- | --- | --- | --- |
| **All episodes** |  |  |  |  |
| any evidence of<br>symptoms | 9476 (49%) | **2880 (15%)** | 4602 (24%) | 2283 (12%) |
| fatigue weakness | 14369 (73%) | **1667 (8%)** | 3027 (15%) | 642 (3%) |
| headache | 14731 (75%) | **1347 (7%)** | 3084 (16%) | 543 (3%) |
| cough | 14570 (74%) | **1262 (6%)** | 3143 (16%) | 730 (4%) |
| loss of taste | 16474 (84%) | **1113 (6%)** | 1802 (9%) | 316 (2%) |
| loss of smell | 16579 (84%) | **1080 (5%)** | 1710 (9%) | 336 (2%) |
| muscle ache myalgia | 15875 (81%) | **1077 (5%)** | 2395 (12%) | 358 (2%) |
| sore throat | 16370 (83%) | **887 (5%)** | 2138 (11%) | 310 (2%) |
| shortness of breath | 17318 (88%) | **887 (5%)** | 1248 (6%) | 252 (1%) |
| fever | 16690 (85%) | **678 (3%)** | 2100 (11%) | 237 (1%) |
| nausea vomiting | 18135 (92%) | **496 (3%)** | 966 (5%) | 108 (1%) |
| diarrhoea | 18464 (94%) | **412 (2%)** | 764 (4%) | 65 (0%) |
| abdominal pain | 18552 (94%) | **376 (2%)** | 718 (4%) | 59 (0%) |
| **Restricting to episodes with at least 3 visits within 35 days, including the index positive** |  |  |  |  |
| any evidence of<br>symptoms | 2132 (39%) | **1759 (32%)** | 486 (9%) | 1131 (21%) |
| fatigue weakness | 4009 (69%) | **1044 (18%)** | 395 (7%) | 341 (6%) |
| headache | 4120 (71%) | **914 (16%)** | 425 (7%) | 330 (6%) |
| cough | 4160 (72%) | **819 (14%)** | 392 (7%) | 418 (7%) |
| loss of taste | 4591 (79%) | **760 (13%)** | 246 (4%) | 192 (3%) |
| muscle ache myalgia | 4488 (78%) | **731 (13%)** | 348 (6%) | 222 (4%) |
| loss of smell | 4660 (80%) | **714 (12%)** | 228 (4%) | 187 (3%) |
| sore throat | 4641 (80%) | **621 (11%)** | 316 (5%) | 211 (4%) |
| shortness of breath | 4950 (86%) | **554 (10%)** | 158 (3%) | 127 (2%) |
| fever | 4826 (83%) | **488 (8%)** | 305 (5%) | 170 (3%) |
| nausea vomiting | 5247 (91%) | **335 (6%)** | 131 (2%) | 76 (1%) |
| diarrhoea | 5371 (93%) | **261 (5%)** | 112 (2%) | 45 (1%) |
| abdominal pain | 5416 (94%) | **227 (4%)** | 106 (2%) | 40 (1%) |

